# School and community reopening during the COVID-19 pandemic: a mathematical modeling study

**DOI:** 10.1101/2021.01.13.21249753

**Authors:** Pei Yuan, Elena Aruffo, Nick Ogden, Yi Tan, Evgenia Gatov, Effie Gournis, Sarah Collier, Qi Li, Iain Moyles, Nasri Bouchra, Huaiping Zhu

## Abstract

**Background:** The closure of communities, including schools, has been adopted to control the coronavirus disease 2019 (COVID-19) epidemic in most countries. Operating schools safely during the pandemic requires a balance between health risks and the need for in-person learning. We use compartmental models to explore school reopening scenarios.

**Methods:** Using demographic and epidemiological data between July 31 and November 23, 2020 from the city of Toronto, we developed a Susceptible-Exposed-Asymptomatic-Infectious-Recovered-Hospitalized-Isolated model. Our model with age, household, and community transmission allow us to study the impact of schools open in September 2020. The model mimics the transmission in households, the community, and schools, accounting for differences in infectiousness between adults and children and youth and adults’ working status. We assessed the extent to which school opening may have contributed to COVID-19 resurgence in the fall and simulated scenarios for the safe reopening of schools up to May 31, 2021. We further considered the impact of the introduction of the new variant of concern.

**Findings:** Though a slight increase in infections among adults (2.8%) and children (5.4%) is anticipated by the end of the year, safe school opening is possible with stringent nonpharmaceutical interventions (NPIs) decreasing the risk of transmission in the community and the household. We found that while school reopening was not the key driver in virus resurgence, but rather it was community spread that determined the outbreak trajectory, brief school closures did reduce infections when transmission risk within the home was low. When considered possible cross-infection amongst households, communities, and schools, we found that home transmission was crucial for mitigating the epidemic and safely operating schools. Simulating the introduction of a new strain with higher infectiousness, we observed substantial increases in infections, even when both schools and communities are closed.

**Interpretation:** Schools can open safely under strict maintenance of strict public health measures in the community. The gradual opening of schools and communities can only be achieved by maintaining NPIs and mitigating household transmission risk to avoid the broader escape of infections acquired in schools into the community via households. If the new COVID-19 strain is more infectious for children, public spaces, including schools, should be closed, and additional NPIs, including the use of masks, should be extended to toddlers.

**Funding:** This research was supported by Canadian Institutes of Health Research (CIHR), Natural Sciences and Engineering Research Council of Canada, and York University Research Chair program.

**Research in context:** *Evidence before this study:* The design of a gradual school reopening strategy remains at the heart of decision-making on reopening after shut-downs to control the epidemic. Although available studies have assessed the risk of school reopening by modelling the transmission across schools and communities, it remains unclear whether the risk is due to increased transmission in adults or children and youth.We used GoogleScholar and PubMed searches to identify previous published works. We used te following terms: “school closure”, “covid 19 school closure”, “reopening schools”, “reopening screening school”, “school household second wave model”. The search of the studies ended in January 2021. Papers in other languages than English and letters were excluded from the search. Two modelling studies examined the effects of screening and delayed school reopening, two other agent-based modelling studies explored the epidemic spread across different age groups.

*Added-value of this study:* We find that the resurgence of COVID-19 in Toronto in fall 2020 mainly resulted from the increase of contact rate among adults in the community, and that the degree of in-person attendance had the most significant impact on transmission in schools. To our knowledge, our work is the first to investigate the resurgence in infections following school reopening and the impact of risk mitigation measures in schools operation during the pandemic. Our novel and comprehensive model considers the age and household structure, but also considers three different settings, school, household and community. We further examined the effects of self-screening procedures, class size, and schooling days on transmission, which enabled us to compare scenarios of school reopening separately for both adults and children and youth, and model the cross-infection between them to avoid potential underestimation. We found that after schools opened, reducing household transmission was crucial for mitigating the epidemic since it can reduce cross-infection amongst households, communities and schools. Lastly, given the recent report of SARS-CoV-2 variant (VOC202012/01), we investigated the impact of the new variant that may be more infectious in children and youth.

*Implications of all the available evidence:* Our analysis can inform policymakers of planning the safe reopening of schools during COVID-19. We suggest that integrating strict NPIs and school control measures are crucial for safe reopening. When schools are open, reducing transmission risk at home and community is paramount in curbing the spread of COVID-19. Lastly, if children are more susceptible to the new COVID-19 VOC, both schools and community must be closed, the time children spend in essential services locations minimized, and NPI’s for those aged less than three years enforced.

## 1. Background introduction

Education has been severely disrupted by the COVID-19 pandemic. In most countries, the epidemic was controlled in spring 2020 by restrictive measures such as travel bans and closures of non-essential businesses and educational establishments. By mid-April 2020, 94% of learners worldwide were affected by the pandemic, representing 1.58 billion children and youth (C&Y), from pre-primary to higher education, in 200 countries^1^. Although school closure may help control the epidemic^2^, it results in significant detrimental effects, including affecting children’s learning, placing a high burden on the parents, and reducing economic productivity^3^. Hence, policymakers worldwide have had to make difficult decisions about whether and how to reopen schools over the past several months. To date, there has been no easy answer or single standard^4^.

Several studies have highlighted that to maintain control of the epidemic as we ease restrictive closures, stringent nonpharmaceutical interventions (NPIs) need to be in place. These include effective and rapid case detection with isolation of cases, effective and rapid contact tracing and quarantine of contacts, and maintenance of distancing by the public^5-7^. Many countries reopened schools over summer 2020 and beginning of September, adapting multiple control measures, such as cohort approach, distantiated single desks, use of masks, and hand sanitizer^8,9^. However, the cases in schools have been reported continuously^10^. Risk mitigation measures in schools, such as daily self-screening for symptoms and virtual attendance^11^, may help minimize infections in school-aged children, and subsequent transmission of infections acquired in schools into the wider community. Still, the effectiveness of these and other mitigation strategies have yet to be examined.

The efficiency of school closure to mitigate infectious disease for different diseases, including COVID-19, has been widely studied^12-18^. However, school closure is not a long-term solution, so schools’ safe reopening is a crucial issue. Few studies have built compartmental models to examine the potential effects of school opening mitigation measures as the epidemic progressed^18,19^. For example, Paltiel^18^found that screening every 2 days using a rapid, inexpensive, and even poorly sensitive (>70%) test, coupled with strict behavioral rules, was estimated to maintain a controllable number of COVID-19 infections and permit the safe return of students to campuses. Others studies using agent-based models also suggested school reopening may lead to a second wave without strengthening NPIs^3,20^.

While these studies have highlighted the important role of school reopening in the trajectory of COVID-19, the transmission of infections acquired in schools into the wider community via transmission in households with children has not been considered. Furthermore, to date there has been no literature on whether the resurgence of COVID-19 was driven by increasing of contacts between C&Y or adults. In December 2020 a new COVID-19 variant of concern (VOC) in UK was reported^21^ and, the increasing number of cases in ages 0-19 years maybe that C&Y are more susceptible to the variant^22,23^. Consequently, school reopening after the holiday is still under discussion. Although there is currently no evidence regarding this new strain’s infectiousness, it is critical to examine the potential impact of higher susceptibility in schooling children for a safe school reopening strategy.

We developed a compartmental model that considers age groups, household structure and transmission, coupled with different levels of population-wide NPIs and school mitigation strategies, to explore potential scenarios for the safe reopening of schools following the winter break. We used demographic and epidemiological data from the city of Toronto, Canada, across the multiple phases of escalation and de-escalation policies regarding schools and community. The model framework, analyses and conclusions can be easily applied to other regions.

## 2. Method

### 2.1 Model description and assumptions

We explore the impact of school opening using a deterministic age-household-location-structured Susceptible *S*-Exposed *E*-Asymptomatic *A*-(subclinical) – Infectious *I*_2_(prodromal phase)-Infectious *I*_2_ (with symptoms) - Recovered R model framework, including two further compartments, depending on infection severity: hospitalization (H) and fully isolated (W) (SEAIRHW). The population is classified into adults (>19) and children and youth (0-19), which labeled as *a,c* respectively. The location structure includes households, schools, and community. Using census data of Toronto in 2016^24^, we established the average household size of 2 for households without children, and 3 for households with children. Depending on the working status of the adults, we further classify the household into working-from-home (in subscript q), with no social activity, and working-outside-of-home (in subscript g), with social activities in the community. When schools reopened, families were able to opt for either in-person or virtual learning. Hence, a proportion of C&Y attends school in person (in subscript sc) or stays at home (in superscript h). For notational convenience, henceforth we will indicate the working-from-home household with ‘WFH’ and household working outside the home with ‘WO’. Population and community classifications are reported in Appendix Table A1.

We model transmission over two periods: before and after schools reopened in September 2020. Before school reopened, all individuals from WFH are assumed to be susceptible, but, once C&Y go back to school, adults may return to work. This will convert these households from WFH to WO. While schools are closed, C&Y are at risk of infection in their household. If C&Y are also part of a WO (socially active) household, the risk of infection will be possible from both family members and the community. After schools reopen, C&Y attending school in person will further face risk of infection from within the school. Similarly, adults can be infected by household members including children, or, if belonging to WO, from the community as well.

NPIs have been shown to be extremely important for controling the epidemic. We include detection and isolation of COVID-19 cases by testing and tracing, implementation of quarantine at home of people contacting infected cases, as well as social distancing^25^. Also, control measures specific to the school setting included: quarantine and contact tracing of school cases, mandatory self-screening procedures, optional in-person attendance. Figure 1 describes the dynamics of our model. See Appendix A for details on returning (back-to-school) rate, going out rate, quarantine, isolation ratios, household structure and model equations (see Appendix Tables A2-A3 for model assumptions, variables, and parameters).

**Figure 1:**
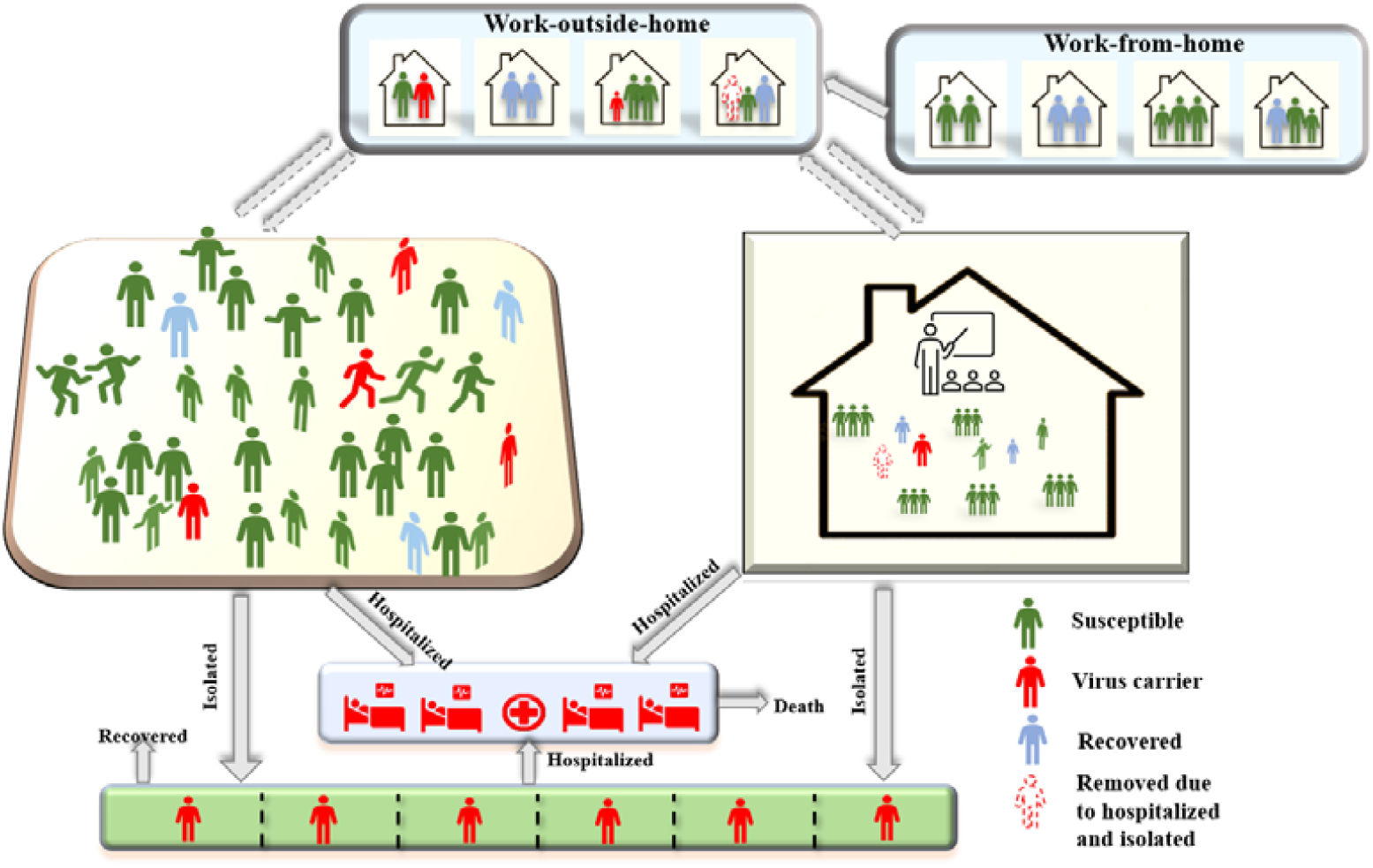
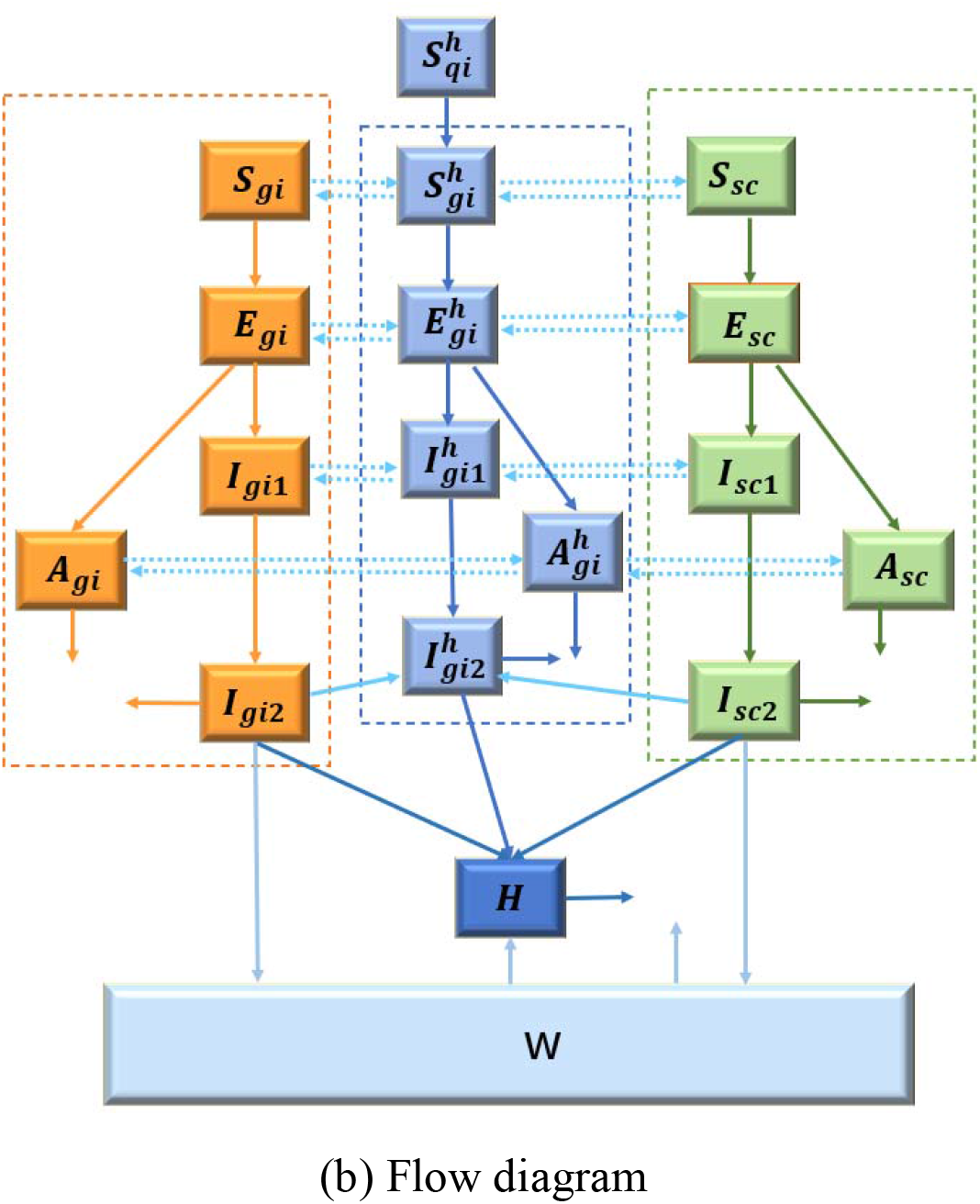
Modeling with age and household structure. (a) shows the activity and response of different groups. (b) Schematic diagram, solid lines indicate movement between classes. *i* indicates the different class of population (*i*= *a,c*).

The instantaneous reproduction number (*R_t_*) is estimated by using daily new cases by episode date^26,27^, assuming a Gamma distributed serial interval range from 2.2-7.8 days with a standard deviation of 4.1-5.9 days considering the serial interval is shorten over time by NPIs^28^. Also, we estimate *_t_* on weekly sliding windows.

### 2.2 Data sources and calibration

According to census data, the average household size in Toronto is 2.4, and 38.39% have children^24^. For simplicity, we assume that all the households have 2 or 3 members, depending on whether they have children. Our model is calibrated using publicly available surveillance data^26^. The optimal values of the per-contact transmission probability of adults, the average number of contacts among adults in the community, the transmission risk in the household, the quarantine rate of symptomatic adults and C&Y, the maximum rate of the adult from WFH back to work and efficiency of self-screening procedures are estimated by minimizing the sum of squared differences between the model’s estimates of adults and C&Y confirmed case and data on these two indicators between Jul 31, 2020, and Nov 23, 2020, collected from the Toronto government website^26^. The parameters estimated and from the literature are presented in Table A4 (Appendix A).

Our estimation shows that the per-contact transmission probability increased by 8.6% (from 2.41% to 2.62%), and up to 25.7% of adults returned to work after the school opened. The contact rate between adults rose by 11.7%, and the contact rate between adults and children rose by 9.1%. Surprisingly, the contact rate for children only increased by 1.5%.

## 3. Results

### 3.1 The risk of school reopening

Our results show that opening schools will lead to more infections in C&Y than in adults (figure 2A,B). However, if the contact rate in the community and per-contact transmission probability remains unchanged (i.e., current transmission risk, as of the date of Nov 23) after schools opened, that is, people adhere to social distancing and other NPIs are in place, the school reopening may not cause a large increase in infections (2.5% increase in adults and 5.4% increase in C&Y by Dec 31, 2020, figure 2A,B). Similar trends are shown under low transmission risk if public health control measures are strengthened (0.8% increase in adults infections and 2.6% increase in C&Y infections by Dec 31, 2020, figure 2C,D).

**Figure 2:**
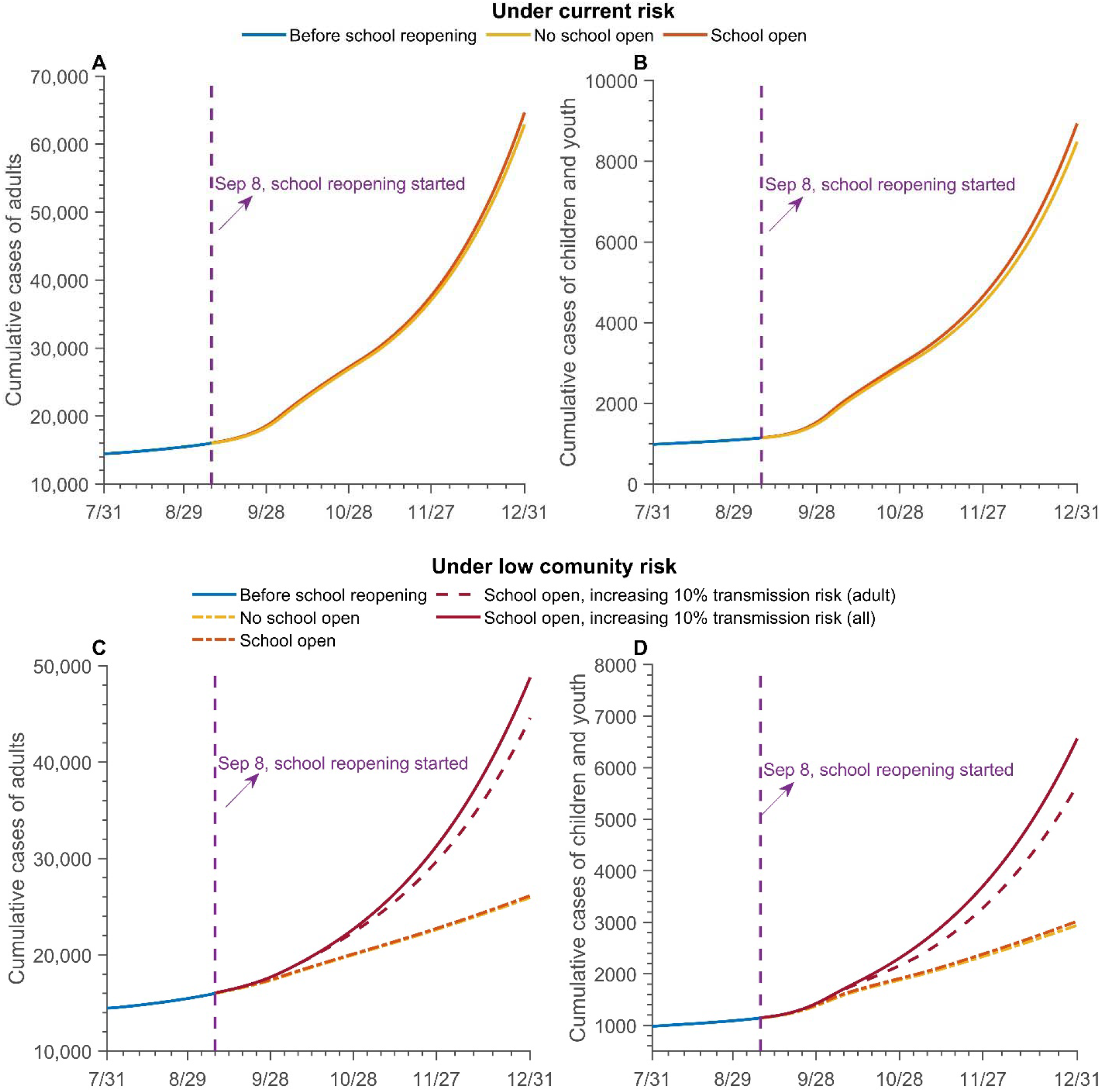
Different scenarios of cumulative cases of adults, children, and youth under current and low risk. The risk levels are referred to the contact transmission risk in the community. (A-B) The current risk refers to the model estimation. (C-D) The low community transmission risk refers to *β*_*a*_ = 1.9%, *c*_*aa*_ = 9.429 after school reopening. Current transmission risk is the estimates as the date of Nov 23. *β*_*a*_ is the per-contact transmission probability. *c*_*aa*_ is the contact rate between adults in the community. Sep 8 is the date of schools starting to reopen, and C&Y are gradually back to school after that. Taking into account the timely adjustment of public health control measures, when studying the impact of school opening on the epidemic, we only simulated until the end of December. Extending the simulation time shows the same trend.

On the other hand, if the contact rate in the community increases by 10% after schools open, the cumulative infection of adults and C&Y will increase significantly (88.0% and 123.1% by Dec 31, 2020, respectively, figure 2C,D). If only the infection rate between adults in the community increases by 10%, this results in a large increase in infections among adults and C&Y (71.8%, 92.4% by Dec 31, 2020, respectively, figure 2C,D). This may give us some insight that the secondary outbreak after the school opened may result from the increase in the contact rate between adults in the community.

### 3.2 Optimal safe reopening strategies to mitigate the current rising trend

Under current risks, the *R*_*t*_ of Toronto on Dec 31, 2020 calculated based on model was around1.2. Given the current contact rate in the community, per-contact transmission probability and the home transmission risk are too high to keep the instantaneous reproduction number (*R*_*t*_) below 1. To control the *R*_*t*_ below 1, the risk of home transmission needs to be reduced to 0.5%, and the contact rate between adults in the community needs to be kept below 9, if the current adult’s per-contact transmission probability is 2.19% (1.1% for children)(figure 3A,B). Also, suppose the current community contact rate is maintained. In that case, the home transmission risk needs be reduced to 0.5% to control the epidemic, although the per-contact transmission probability reduced to 1.8% (0.9% for children)(figure 3C,D). After schools are open, controlling the transmission risk at home and in the community is crucial to mitigate the epidemic.

**Figure 3:**
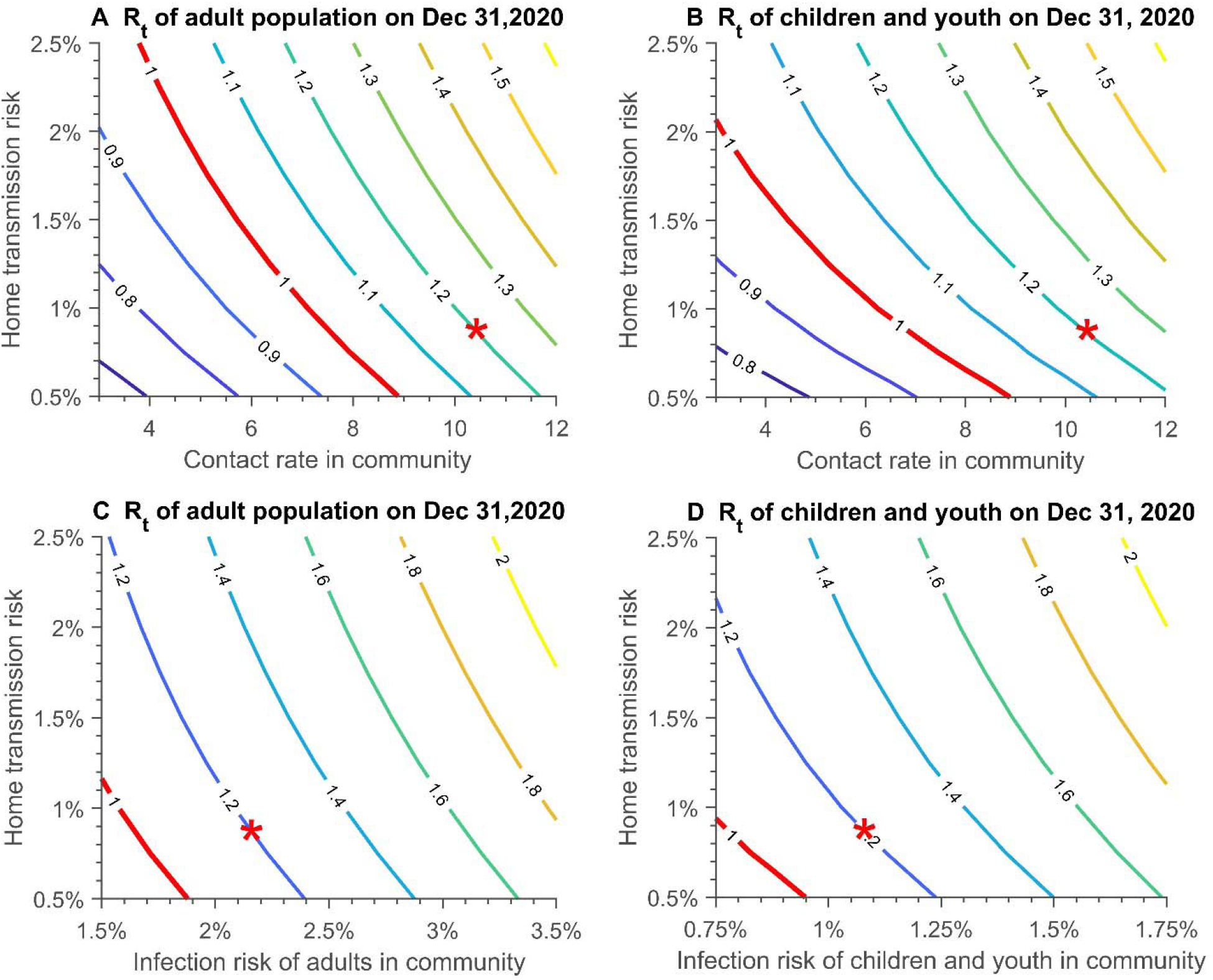
Instantaneous reproduction number (*R*_*t*_) on Dec 31, with varying household and community transmission risk. (A) (C) Adult population; (B) (D) Children and youth population. Red star represents the current state. The contact rate in the community refers to the contact rate between adults. Infection risk is the per-contact transmission probability.

### 3.3 Modeling predictions

In the short term, school closures for a period (extending the holiday period, assumed two weeks in the simulations), can reduce the spread of the epidemic to a certain extent (figures 4A, B), on the premise that the risk of family transmission does not rise in the holiday period. The least risky scenario is when extending school closures and the contact rate in the community is reduced by 10%, and the cumulative infections of adults and C&Y by Jan 31 will drop by 26.6% and 25.2%, respectively. However, the worst case will happen if the household transmission risk increases by 10% during the period of school closures, and the extended vacation will increase the spread of the epidemic in both the adult population and the C&Y population (2.6% increase in adults and 3.9% increase in C&Y by Jan 31, 2020).

**Figure 4:**
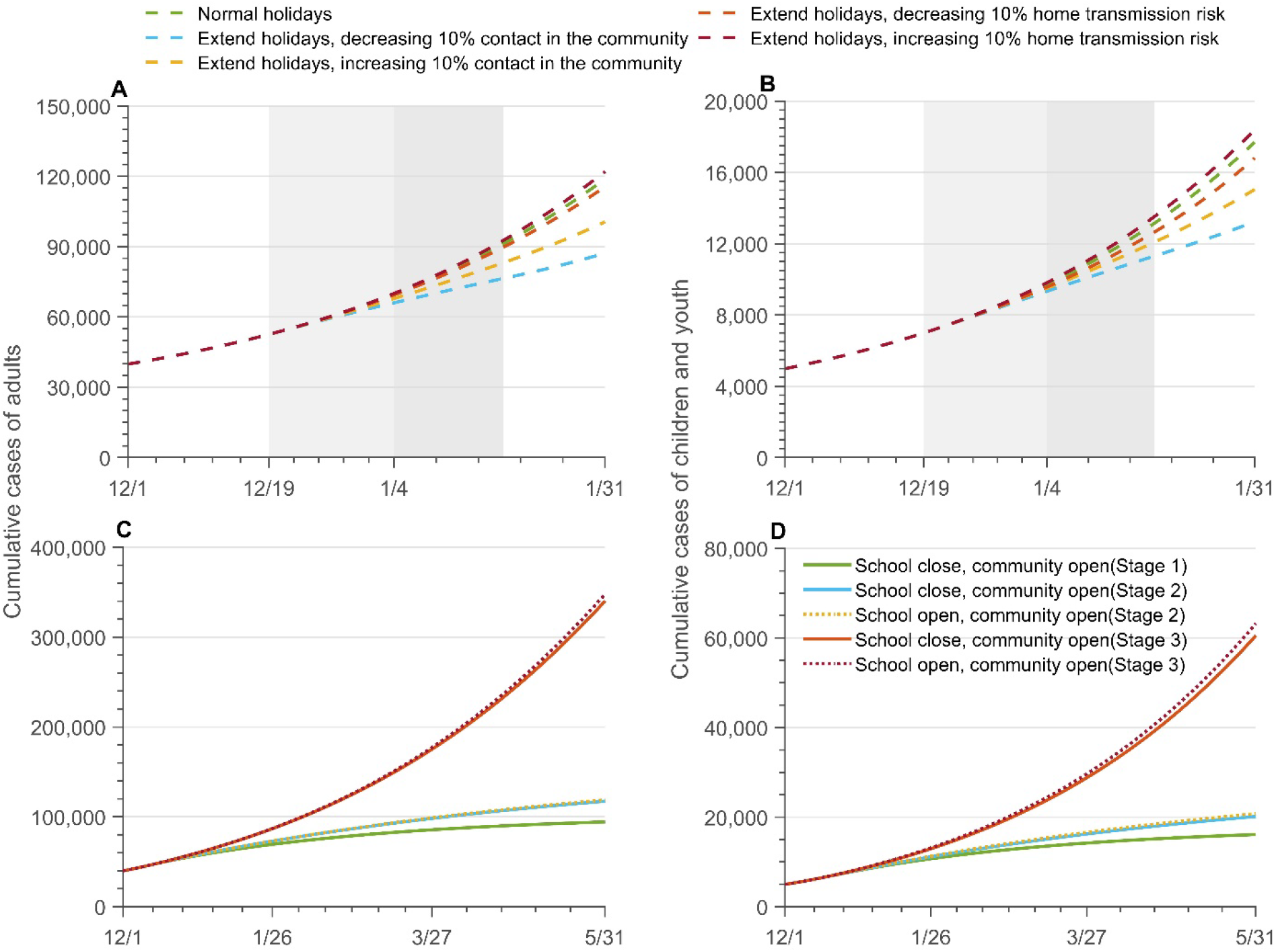
The projection of cumulative cases with different senarioes. Forecast of cumulative cases of (A) adult and (B) children and youth infections till Jan 31, 2021 under different transmission risk when extend the holidays two weeks; The projection of cumulative cases in adults (C), and children and youth (D) up to May 31, 2021 under different opening strategies. The dot lines refer to the school opens while solid line represents the school close. School opens with strictest NPIs, and the community opens in different levels (like the reopening stage during May to Sep in Toronto, the detailed information about stages are included in the appendix).

When extending the forecast to May 31, 2021, our model predicts that school reopening is not the key to the outbreak of the epidemic, but the risk of transmission in the community determines the trend of the epidemic (figures 4C, D). If the community risk is low, safe school opening is feasible, when strict NPIs measures are implemented and the community transmission risk is not increased after the school opens. Although the infection of adults and C&Y will increase slightly, and the increment of C&Y infection will be higher than adults after the school opening. However, The stricter NPIs and school closures may be needed if the new strain is introduced which has higher transmission probability and more infectious for C&Y. The new variant may cause a large outbreak even under the strictest control measures(the number of infections of adults and C&Y till May 31, 2021 under the new strain virus is 3.6 times and 4.2 times that of the old strain, figures A4). An exponential increasing epidemic occurs irrespective of whether schools are closed or open with high community transmission risk due to weak NPIs (community is fully opened as in stage 3 in Ontario, Table A5).

## 4 Conclusion and discussion

Our novel and the complex model considers age structure and household transmission, allowing us to examine three different risks within and between households, communities, and schools, and to explore is the school reopening responsible for the fall outbreaks. We found that school opening can be safe under strict NPIs, although there will be a slight increase in infections among adults (by Dec 31, 2.8%) and children (5.4%). However, whether the reopening of schools can cause another outbreak mainly depends on the countermeasures in the community. The increase in cases occurring when schools are opened is primarily due to the rise of contact between adults who can now return to work and social activities. Overall, reducing the contact rate in the community is more effective in mitigating the epidemic. Nonetheless, reducing the contact rate in the community has a larger impact on reducing the cases of adults, while reducing transmission risk within the home is more effective in reducing infections in C&Y.

The safe operations of schools during the pandemic necessitates risk mitigation measures. The effectiveness of those measures is examined (figureA3, table A6). Attendance is the biggest driver of infection; with high attendance, we also need these extra measures that are in place in Toronto. Also, to enable the safe opening of schools, it is not sufficient to control the spread in the community. When schools open, the increase in adults and C&Y activities, and the risk of transmission within the household will lead to an increase in cross-infection risk in schools, communities, and households. Hence, controlling household transmission risk is paramount. Only at low levels of risk of family transmission can schools and communities be open, given a similar overall infection risk. Our predictions show that, in the long term, if the household transmission is not reduced, the cases will keep increasing even if schools remain closed for 2 additional weeks following the holiday break. Though reducing household transmission is difficult, public health communication should emphasize preventative practices for students returning from school, including washing hands, changing clothes, and mask-wearing. Besides, the aggressive testing, tracing, isolation of mild cases^29^ (figure A5), the Fangcang shelter hospital^30^ both are effective to decrease the spreading, then to reduce the infection in the household. Nevertheless, the key to allowing safe school opening is the maintenance of strict NPIs usage in the community and reducing community spread.

We also examined the consquences of the new COVID-19 variant of concern. Even with strict NPI’s and school closures, the number of cases could rise rapidly within a few months. Our findings suggest that assuming that C&Y are more susceptible to the new virus variant, the city should immediately shut down the communities and schools. Moreover, if the new strain is more infectious in younger children less than 3 years of age, reducing the amount of time they spend in the community and increasing their use of NPIs, such as mask-wearing, should be considered.

Our analyses have some limitations. Firstly, our school compartment does not include adults, such as teachers, employees, and staff working in the school. However, in the community, we include all three types of contacts between adults and C&Y. When schools are opened, the contact between adults and C&Y in schools can be reflected in the increase of that in the community. Therefore, this reasonable simplification can also fit our purpose within this analysis. Secondly, the age classification in our model only includes adults and C&Y, and no more detailed classification is added, hence the vulnerable person was not included in this study. Due to the incorporation of the household structure, the complexity of our model has significantly increased, the overall model dimension has risen more than 300. In addition, the results of our model have well explained the impact of school opening on the epidemic. Our reasonable simplification of the age structure will not affect our analysis of school opening risks and prevention and control and our model assumptions have been checked with policymakers. When these measures cannot be effectively implemented, the risk of school opening needs to be reconsidered and evaluated. Also, in the long-term forecast, our assumptions about the contact rate in the community and the per-contact transmission probability may be too optimistic. The results of long-term forecasts should be considered as the minimum risk. At the same time, it did not consider that the government might adopt other control measures after the infection increased. We are more concerned about the comparison of the impact of different levels of openness of schools and communities on the development of the epidemic. Furthermore, the infectiousness among C&Y aged younger than 19 years is assumed to be 50% of the adults. We make this assumption based on the currently available research suggesting that the susceptibility of children is lower than that of adults. But it may be different from this ratio with further investigation and we also examine the scenarios with new strain variant if more infectious in C&Y population (figure A4).

In summary, our findings suggest that the combination of stringent public health measures to control transmission in the community, mitigation efforts in schools, and efforts to minimize transmission in the household can allow for the safe reopening of schools. However, when schools open, the increasing contacts among adults in the community will lead to another large-scale surge in the epidemic in the absence of adequate NPIs. Controlling transmission within the household, particularly for students returning home from school, could be key to preventing cross-transmission between the household, the community, and the school with both the schools and community settings open.

## Data Availability

The data used for this study are published by the City of Toronto.

https://open.toronto.ca/dataset/covid-19-cases-in-toronto/

## Article information

### Author Contributions

Research design: H.Z., P.Y., E.A., EvgeniaG., N.O.; Literature search: Q.L., Y.T.,E.A., P.Y.; Data collection: EvgeniaG., P.Y. ,Y.T.,Q.L.; Modeling: H.Z. and all; Model analysis: P.Y., H.Z.; Simulations: P.Y., Y.T.,Q.L.; Draft preparation: P.Y., E.A., Y.T., Q.L., H.Z.; Writing-reviewing-editing: H.Z.,N.O, EvgeniaG., EffieG., S.C., I.M., N.B;Supervision: H.Z.

### Data sharing statements

The data used for this study are published by the City of Toronto, publicly available at the following links: https://open.toronto.ca/dataset/covid-19-cases-in-toronto/.

## Appendix A

### 1. Detailed modeling methodology

On Jul 31, the city of Toronto enters Stage 3 of reopening, nearly all businesses and public spaces can gradually reopen. Also, the school start opening on Sep 8^0^. We build a compartmental model incorporating both age, household, and location structures. The population is classified into adults (>19) and children and youth (0-19), which labeled as *a,c* respectively. The location structure includes households, schools, and community. We consider the following average sizes of households: 2, when the family members are just adults, and 3 for families with 1 child. Also, depending on the working status of the adults, we further classify the household into two categories: working-from-home (in subscript q), with no social activity, and working-outside-of-home (in subscript g), with social activities in the community. After school is reopened, families opt for either in-person or online learning. Hence, a proportion of children and youth is considered to go to school (in subscript sc) or stay at home. For notational convenience, henceforth we will indicate the working-from-home household with WFH and working-outside-home household with WO. The population and community classifications are reported in Table A1. We note that we omit demographic components, such as immigration, birth, and natural death.

**Table A1.** Population, household, and community classifications

|  | WITHIN HOUSEHOLD<br>(in superscript h) |  |  |  | OUTSIDE of HOUSEHOLD |  |
| --- | --- | --- | --- | --- | --- | --- |
|  | WFH (in subscript q) |  | WO (in subscript g) |  | School | Community |
|  | n=2 | n=3 | n=2 | n=3 | (in subscript s) | (in subscript g) |
| **Children and youth** | 0 | 1 | 0 | 1 | Children and youth attending in-person education | Children and youth from WO |
| **Adults** | 2 | 2 | 2 | 2 | 0 | Adults from WO |

A detailed description of dynamical transmission of COVID-19 is described in the flowchart (Fig. 1). Let *N*_*i*_(*t*) (*i*= *g,q*) be the total number of individuals in each sub-group, *g,q,s*, at time Each subpopulation is further the di vided into Susceptible (*S*_*i*_(*t*)), Exposed, (*E*_*i*_(*t*)), Asymptomatic (subclinical) infection (*A*_*i*_(*t*)), Infectious pre-symptomatic (will eventually show symptoms) *I*_*i*1_ (*t*) and Infectious symptomatic (*I*_*i*2_ (*t*)), and Recovered (*R*_*i*_(*t*)). Both and (*A*_*i*_ (*t*)) and *I*_*i*1_ (*t*) are considered to be infectious virus carriers. We assume that individuals in *A*_*i*_ (*t*) will never show symptoms, while individuals in *I*_*i*1_ (*t*) develop into symptomatic classes (*I*_*i*2_ (*t*)) after a specified period of time. Mild symptomatic infections in classes (*I*_*i*2_ (*t*)), may choose to either isolate themselves at home (or other places). If the quarantine is respected well enough, these infections will be fully isolated and, consequently, will not contribute to the spread of the virus. Otherwise, they are still a source of infection until recovery. As the disease progresses, some mild infections may become severe and require hospitalization. We include two further compartments: the fully isolated (*W*(*t*)), and the hospitalized (H(t)) who are all severely affected. It is assumed that neither of these compartments contribute to infection transmission.

Based on the classical SEIR framework, a household-based transmission model with age structure will be proposed to describe the impact of school reopening on the development of the epidemic. Considering that an infected person quarantined at home is interacting only with family members, the number of contacts is limited, so we will use the standard incidence rate in modelling.

Although home transmission is relatively strong, it only involves limited family members. To reflect this, and capture disease transmission within families, we consider the population with households.

For household members from WO, susceptible individuals will (*S*_*qi*_(*t*)) be infected by infectious individuals in the home *A*_*qi*_(*t*), *I*_*qi*1_(*t*), or *I*_*qi*2_(*t*), and in the community. After school reopening, the children and youth will face additional transmission risk from the school. Thus, the adults may face the increase of home transmission risk due to the children and youth back to school. The household from WFH is safe and not involved in the transmission of COVID-19. Additionally, infections that are completely isolated will not be involved in transmission.

#### Before school reopening

The household from WFH: all are susceptible, and isolated. The household from WO:

- Children and youth population (0-19): stay at home, facing the transmission risk from household members and transmission risk in the community.
- Adult population (>19): go to work, or other social activities, facing the infection risk from community, and the risk from household members.

#### After school reopening

The household from WFH: all are susceptible and isolated. The household from WO:

- Children and youth population: Go to school, facing the transmission risk from household members and transmission risk in the community and school.
- Adult population: Go to work (the proportion may increase due to the school reopening), or other social activities, facing the infection risk from community, and the risk from household members.

### 2. Rates definition

#### ⍰ Returning (back to school) rate of children and youth

All the children and youth from WO and proportion of children and youth from WFH will back to school after school reopening. We assume that τ is a random variable which describes how long that the children and youth back to school after school reopening. Hence, τ follows an Exponential distribution

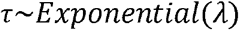

with 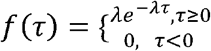, where *λ* is the average number of children and youth back to school every day. The expectation of *τ* = *E* (*τ*) = 1/ *λ* = Δ *T*_*A*_ (Δ *T*_*A*_ is the average completion time for children and youth back to school), and *f*(*τ*) is the probability that children and youth back to school in *τ* days.

The total number of children and youth of Toronto that may back to school at *T*_1_ is *N*_*sc*0 *i*_*= p*_*c*_ *N*_*H*_*\** (1 − *q*_*h*_ + *q*_*h*_\**G*_*q*_), where *p*_*c*_is the proportion of household with children and youth, *N*_*H*_ is the total number of household, *q*_*h*_ is the proportion of WFH household and *G*_*q*_ is the maximum going out rate of WFH (see below for description of going out rate). The number of children and youth who back to school on *T*_1_+*τ* days was Δ*N*_*sc*_ (*T*_1_+*τ*) = *a*_*r*_* *N*_*sc*0_* *f*(*τ*) *a*_*r*_ is the school attendance ratio of children and youth back to school. Let *gA* (*T*_1_+*τ*) be the daily returning rate on day *T*_1_+*τ*, then *g*A (*T*_1_+*τ*) = *a*_*r*_* *f*(*τ*) And it satisfies 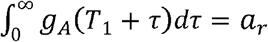 Then the number of children and youth newly back to school on that day is

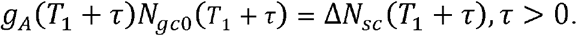

Where. *N*_*gc*0_ (*T*_1_+*τ*) = *S*_*gc*_ (*T*_1_+*τ*) + *E*_*gc*_(*T*_1_+*τ*) + *A*_*gc*_(*T*_1_+*τ*)+ *I*_*gc*1_ (*T*_1_+*τ*).

Hence, we have

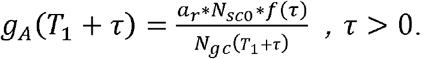

Let *t* = *T*_1_+ *τ*, then

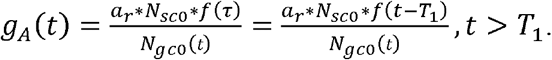

The cumulative returning rate is 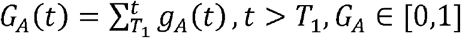.

#### ⍰ Going out rate of WFH household

After school reopening, some adults may return to work due to the children back to school. Hence, the WFH households becomes the WO household. Like the returning rate, we derive the going out rate

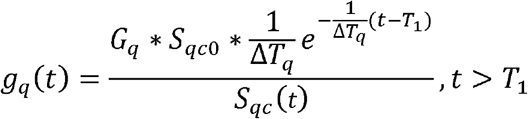

where, *S*_*qc*0_ = *p*_*c*_ *N*_*H*_ *\*q*_*h*_ represents the number of children and youth in the WFH household before the school reopening. Δ*T*_*q*_ is the average completion time for these WFH households that children and youth may back to school. Here, we assume that Δ*T*_*q*_*=* Δ*T*_*A*_=7.

### 1. Model structure and model equations

According to the infection and development process of the disease in the human body, at time t, an individuals in a household can belong to one of the following categories: *S*(*t*), *E*(*t*), *A*(*t*),*I*_1_(*t*) *I*_2_(*t*), *H*(*t*) or *W*(*t*), or maybe recovered *R*(*t*). Corresponding to each disease class, we assign the number of individuals in each household to be *i,j,k,l,m,x,y,z*, respectively and limit households to a size of *n* such that *n*=*i*+*j*+*k*+*l*+*m*+*x*+*y*+*z*. Given the average household size is 2.4 in Toronto, we classify the household into two different type(1−*p*_*c*_) proportion of household without children(*n*=2, 2 adults), and *p*_*c*_ proportion of household with children (*n*=3,2 adults, 1 child). Therefore, each household at most consists of different categories of individuals. Based on the classification and combination of individuals in households, all possible types of households in Toronto are 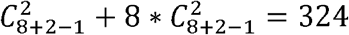. The total population in Toronto is N.

For the WFH household with *q*_*h*_ proportion, we assume they stay at home and individuals are susceptible (*q*_*h*_*[3\**p*_*c*_\**N*_*H*_+2*(1− *p*_*c*_) \**N*_*H*_])and isolated. Since after 6 months epidemic, if they have been infected, they will have shown symptoms or have already recovered. For the WO household, the dynamics are determined by eight processes: within-household transmission; disease progression from Exposed to Asymptomatic infection or Infection without symptoms; disease progression from Infection without symptom to Infected with symptoms; recovery from Asymptomatic infection; recovery from Infected with symptoms; hospitalization of Infected with symptoms; isolation of Infected with symptoms; and transmission in the community. 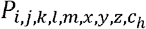 is the number of households with *i* susceptible,*j* exposed, k asymptomatic (subclinical) infection, l infectious without no symptoms, m infected with symptoms, x hospitalized, y isolated and z recovered adults, and with the children *c*_*h*_ at h state, where *h*∈{*S,E,A,I*_1,_ *I*_2,_*H,W,R*,0}. Here, state of 0 means no kids in the households. Then the variation of the number of household with children and youth 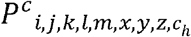 from WO with respect to time *t* (*t*<*T*_1,_ *T*_1_ is the time of school reopening) can be given by

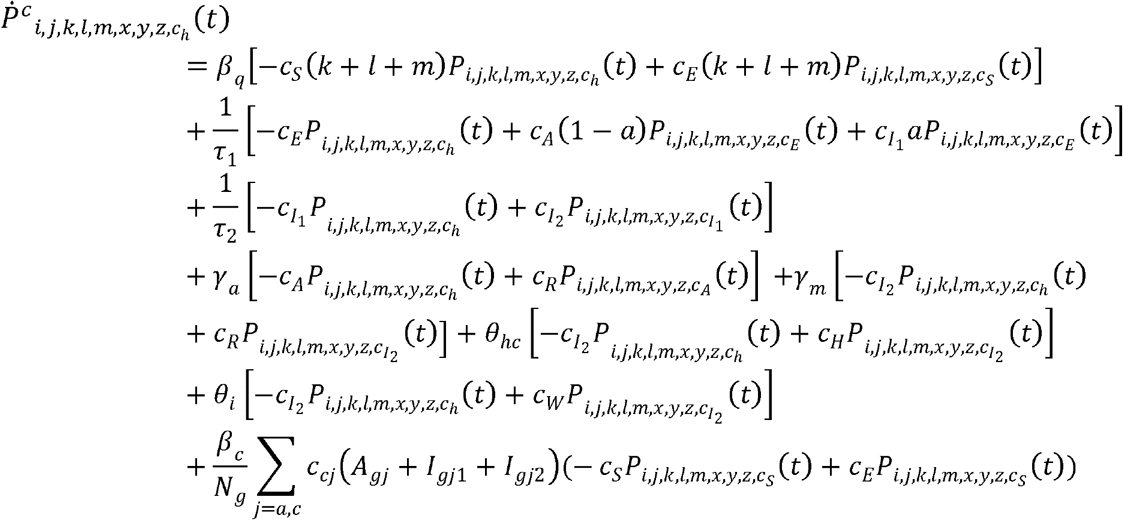

Where

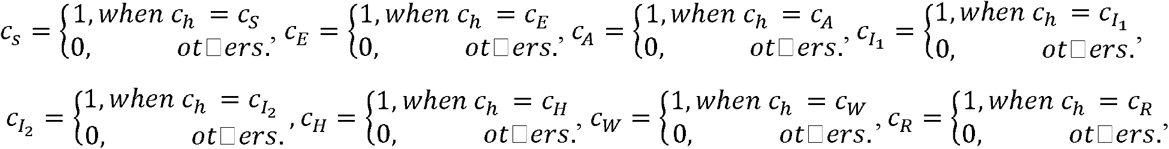

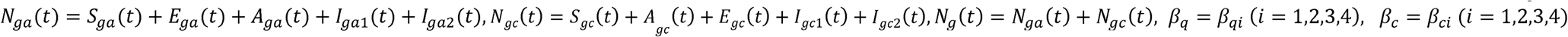 is estimated by different phase based on the data.

Then the variation of the number of households 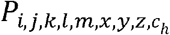 from WO with respect to time *t* (*t*<*T*_1,_ *T*_1_ is the time of school reopening) can be given by

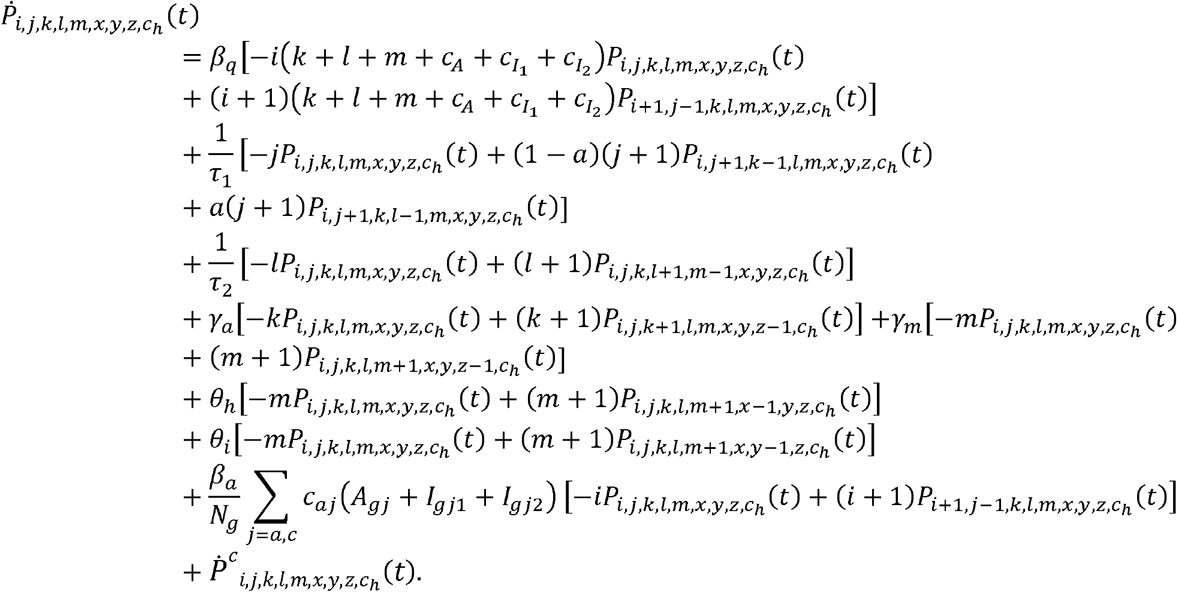

where 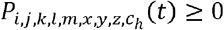 should be satisfied, and *β*_*a*_= *β*_*ai*_(*i*=1,2,3,4) is estimated by different phase based on the data.

#### ✥ Before school reopening *t* < *T*_1_

#### For adult’s population in the community

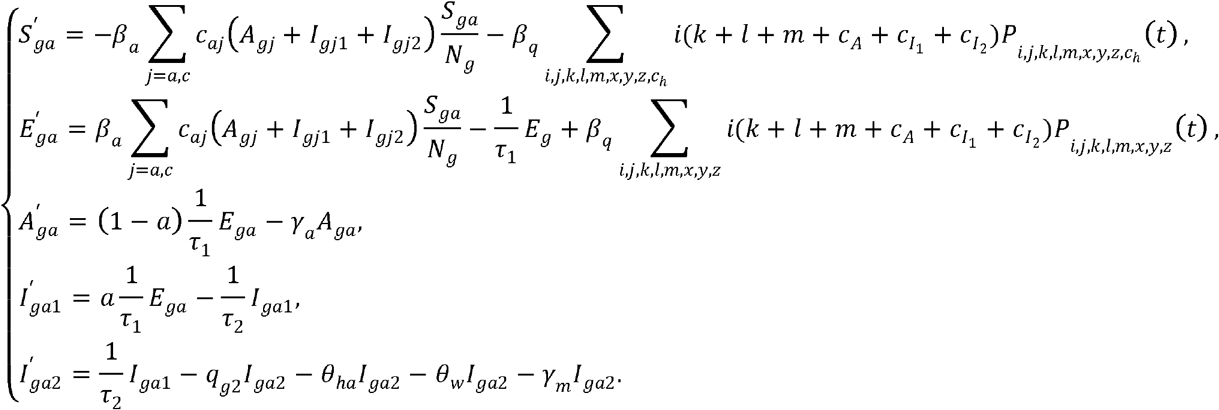

where *c*_*aa*_ = *c*_*aai*,_ *c*_*ac*_ = *c*_*aci*,_(*i*= 1,2,3,4) is estimated by different phase based on the data.

#### For children population in the community

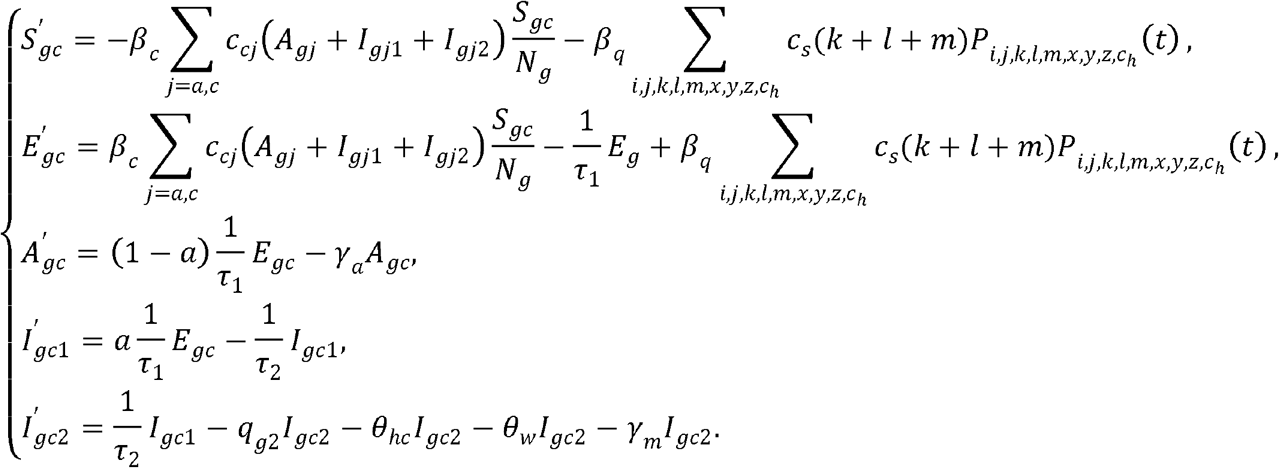

#### For the population in the household from WO

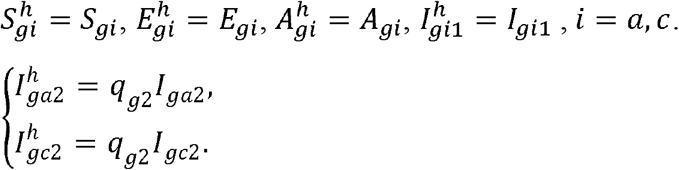

#### For the population from WFH, *t*<*T*_1_

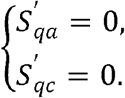

#### The populations of hospitalization, isolation, recovered and deceased

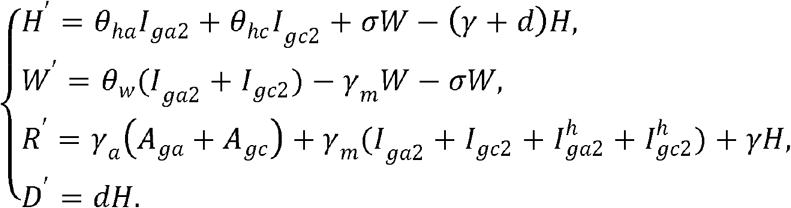

#### ✥ After school reopening

After school reopening, the variation of the number of household with children and youth 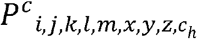 from WO household with respect to time *t* (*t*≥*T*_1,_ *T*_1_ is the time of school reopening) can be given by

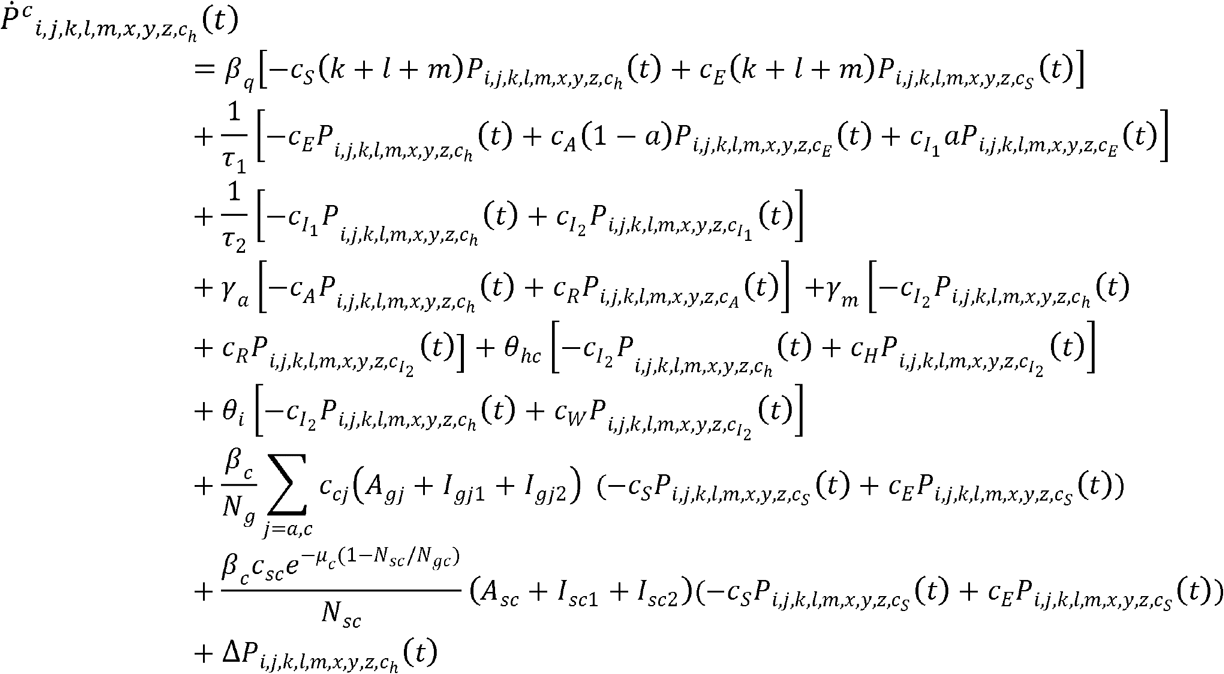

where

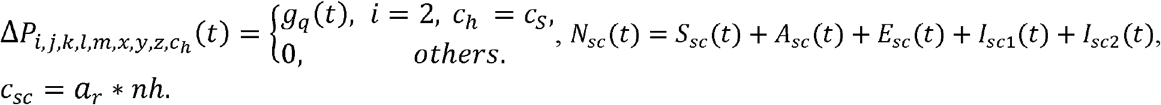

The variation of the number of households 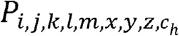 from WO household with respect to time *t* (*t*≥*T*_1,_ *T*_1_) can be given by

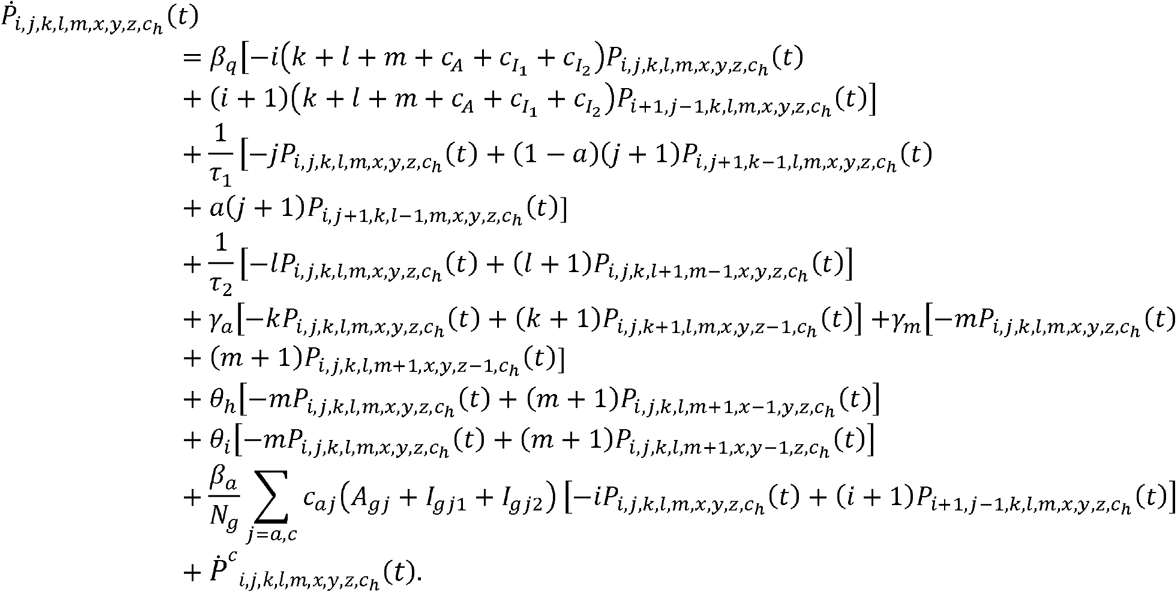

#### For the adult population in the community

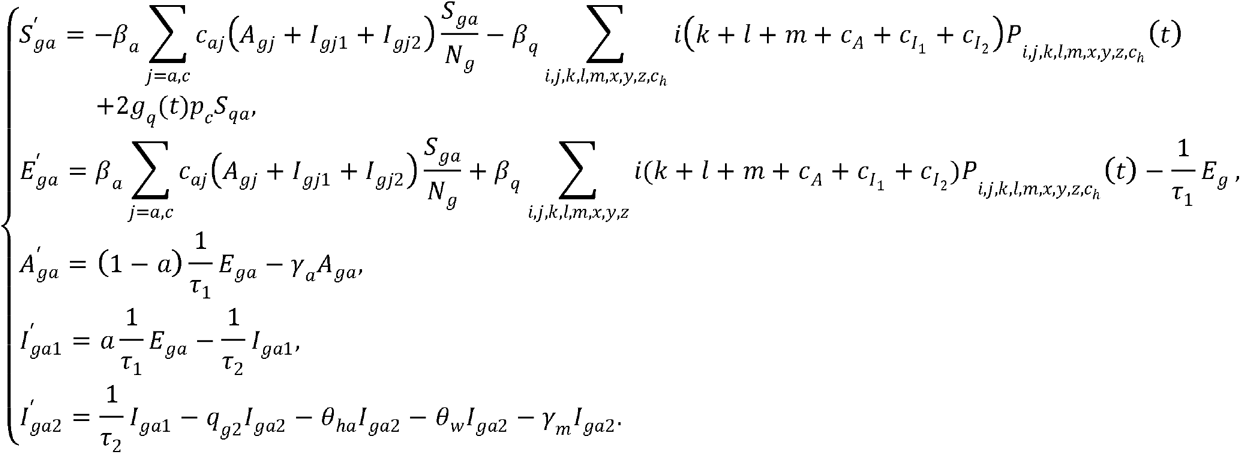

where *c*_*aa*_ = *c*_*aai*,_ *c*_*ac*_ = *c*_*aci*,_(*i*= 1,2,3,4) is estimated by different phase based on the data.

#### For children population in the community

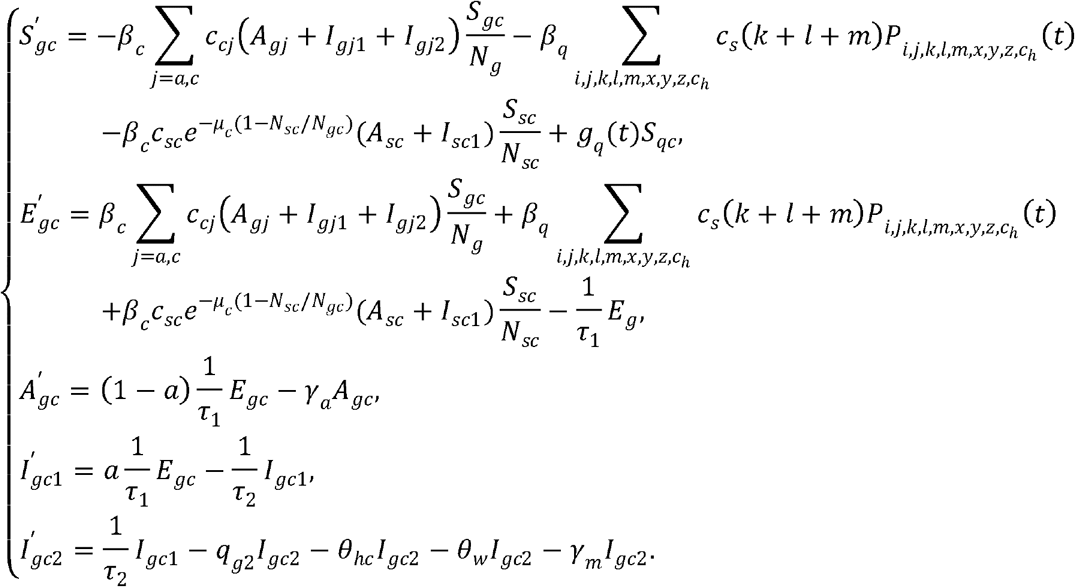

#### For the population in the household from WO

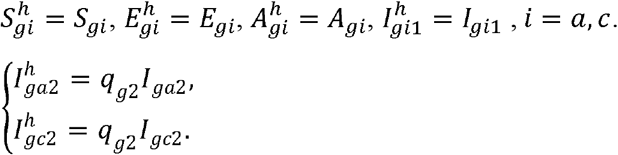

#### Children in the school

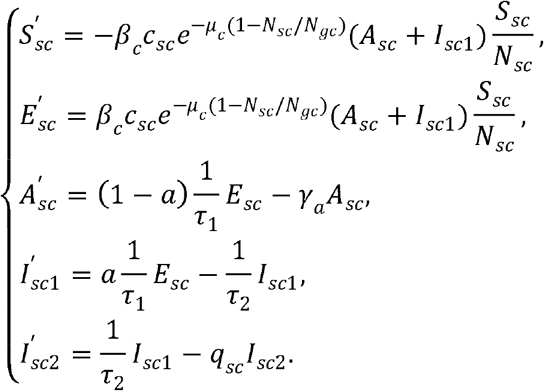

The school opens at 9 am and closes at 3 pm from Monday to Friday.

#### When children go to the school at time *t*_s_

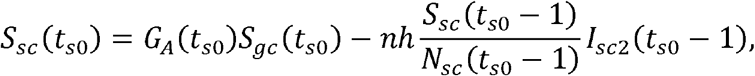

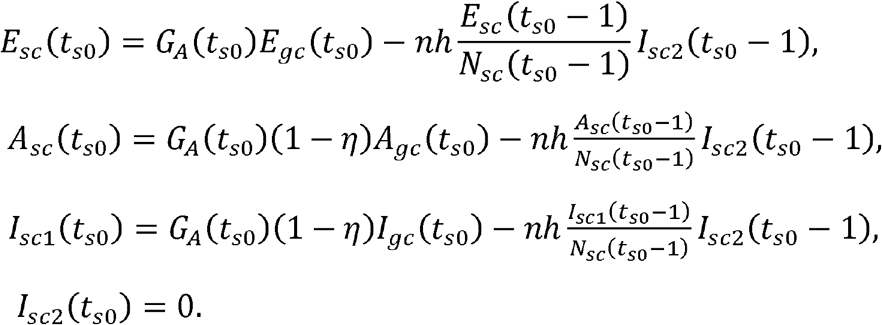

where 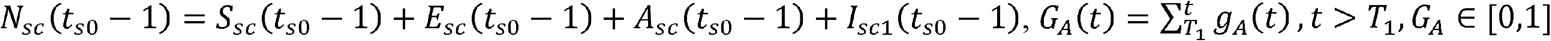

When school close at time t,

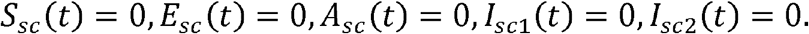

#### For the population from WFH, *t* ≥ *T*_1_

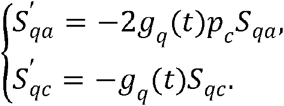

where 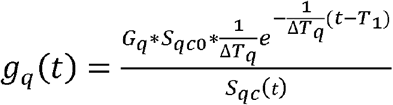

#### The populations of hospitalization, isolation, recovered and deceased

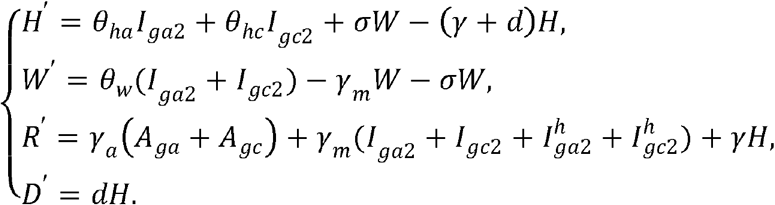

Tables A2-A4 report the assumptions, variables and parameters employed in the study, respectively.

#### The initial value of different household type based on data

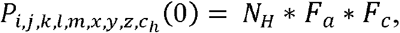

where

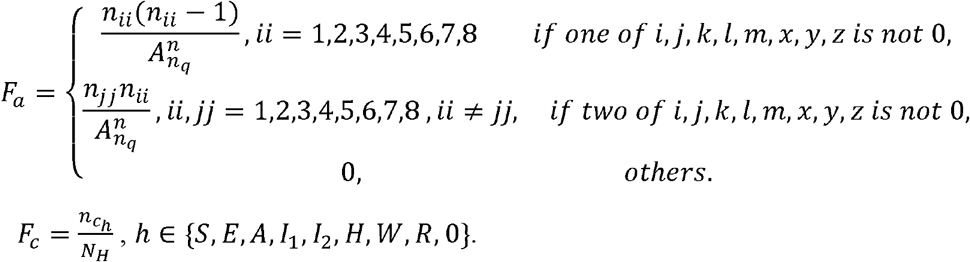

with 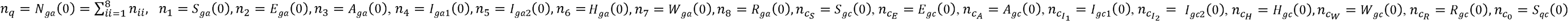

**Table A2.**
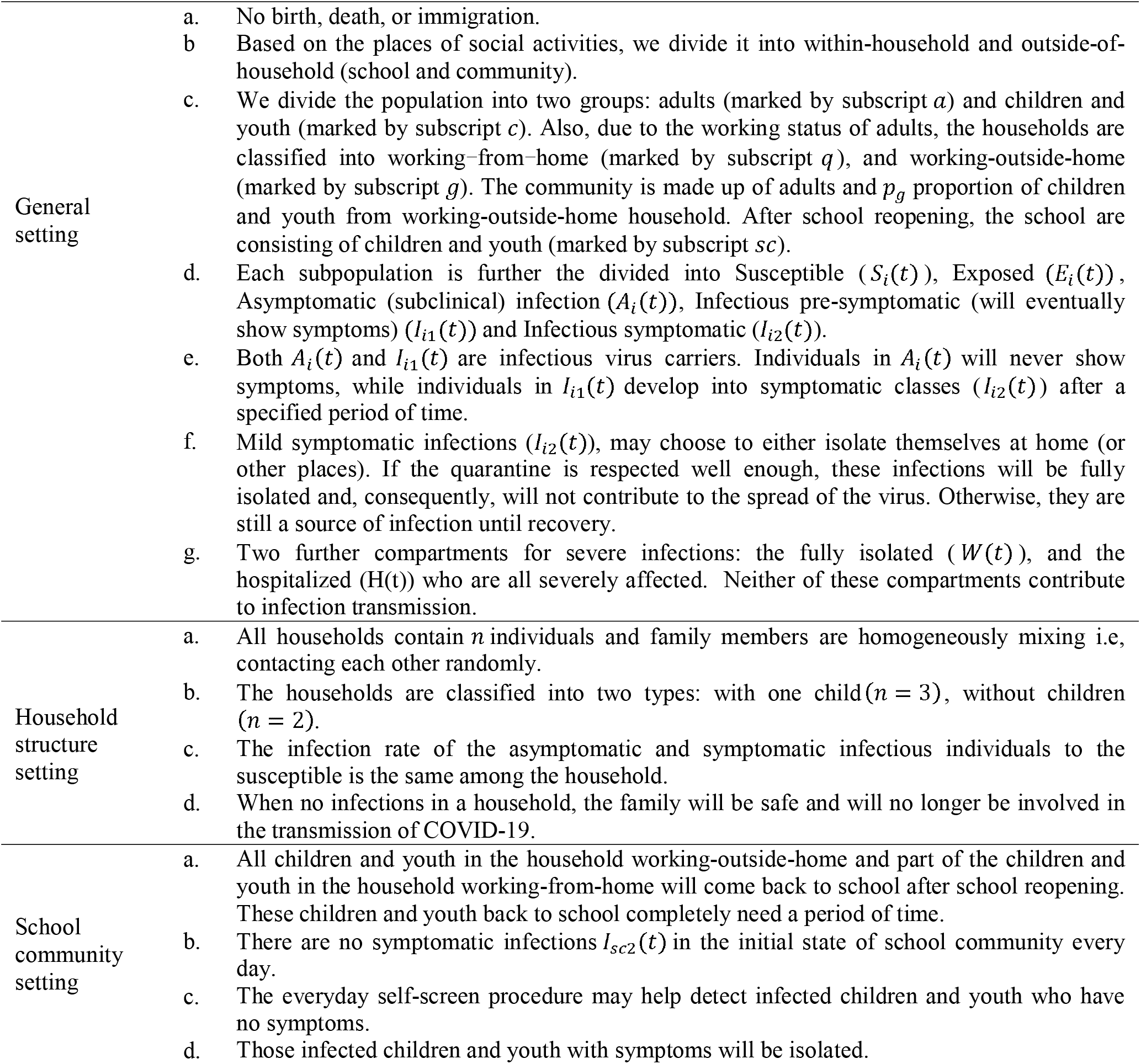
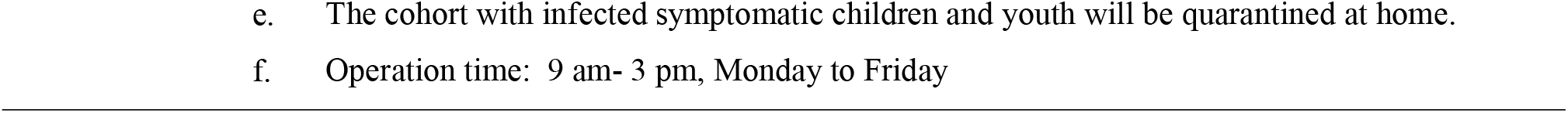
Model assumptions

**Table A3.** Identification of the variables and their initial values

| **Variables** | **Descriptions** | **Fixed initial Values** | **Sources** |
| --- | --- | --- | --- |
| $S_{qa}(t)$ | The number of susceptible adults from WFH household. | 964716 | Calculated |
| $S_{qc}(t)$ | The number of susceptible children and youth from WFH household. | 185177 | Calculated |
| $S_{ga}(t)$ | The number of susceptible adults from WO household. | 1499991 | Calculated |
| $E_{ga}(t)$ | The number of exposed adults from WO household. | 102 | Calculated |
| $A_{ga}(t)$ | The number of asymptomatic infected adults from WO household. | 2 | Calculated |
| $I_{ga1}(t)$ | The number subclinical infected adults from WO household that will become symptomatic | 32 | Calculated |
| $I_{ga2}(t)$ | The number of infected adults with symptoms from WO household. | 296 | 0<sup>[2]</sup> |
| $S_{gc}(t)$ | The number of susceptible children and youth from WO household. | 289607 | Calculated |
| $E_{gc}(t)$ | The number of exposed children and youth from WO household. | 18 | Calculated |
| $A_{gc}(t)$ | The number of asymptomatic infected children and youth from WO household. | 0 | Calculated |
| $I_{gc1}(t)$ | The number subclinical infected children and youth from WO household that will become symptomatic. | 7 | Calculated |
| $I_{gc2}(t)$ | The number of infected children and youth with symptoms from WO household. | 26 | 0<sup>[2]</sup> |
| $I_{sc2}(t)$ | The number of infected children and youth with symptoms in the school. | 0 | |
| $H(t)$ | The number of infections in hospitals. | 84 | 0 |
| $W(t)$ | The number of isolation infections. | 613 | 0 |
| **Variables** | **Descriptions** | **Initial Values** | **Sources** |
| $S_{sc}(t)$ | The number of susceptible children and youth in the school. | - | |
| $E_{sc}(t)$ | The number of exposed children and youth in the school. | - | |
| $A_{sc}(t)$ | The number of inapparent infected children and youth in the school at day t. (that will never develop symptoms) | - | |
| $I_{sc1}(t)$ | The number of infected children and youth without symptoms in the school at day t. (that will become symptomatic) | - | |

**Table A4.** Parameters for COVID-2019 in Toronto, Jul 31-Nov 23,2020

| Parameters | Descriptions | Fixed Values | Sources |
| --- | --- | --- | --- |
| $\tau_1$ | Average time spent in the exposed classes, $E_{ga}$, $E_{gc}$, $E_{sc}$, days | 4 | [3][4] |
| $\tau_2$ | Average time period spent in $I_{ga1}$, $I_{gc1}$, $I_{sc1}$, days | 3 | [4] |
| $a$ | Proportion of infected individuals with apparent infection | 0.953 | [5] |
| $\gamma_a$ | Recovery rate of inapparent infected individuals | 0.07 | [5] |
| $\gamma_m$ | Recovery rate of patients with mild symptoms | 1/14 | [6] |
| $\gamma$ | Recovery rate of patients in hospitals | 0.0357 | [6] (1/42-1/21) |
| $c_0$ | Contact rate before COVID pandemic, 1/day | 11.58 | [7] |
| $c_{ac}$ | The contact rate between adults and children and youth in the community | 4.42 | [8] |
| $c_{cc}$ | The contact rate among children and youth in the community | 8.36 | [8] |
| $\beta_0$ | Probability of transmission per contact among adults in the community | 0.019 | [9] |
| $\beta_{q1}$ | Contact transmission rate within household members | 0.01503 | [9] |
| $c_{ga2}$ | The contact rate among adults outside after school reopening (Jul 31 – Sep 8) | $c_0$ | Assumed |
| $k$ | The factor of the probability of transmission per contact outside the household among children and youth compared to adults | 0.5 | [10][11] |
| $\theta_{ha}$ | Proportion of hospitalization of $I_{ga2}$ | 0.0152 | [9] |
| $\theta_{hc}$ | Proportion of hospitalization of $I_{gc2}$ | 8e-05 | [12] |
| $\theta_i$ | Isolation rate of confirmed cases | 0.039978 | [9] |
| $d$ | Disease-induced death rate in hospitals | 0.034 | [9] |
| $p_c$ | The proportion of household with children and youth | 38.39% | [2] |
| $q_h$ | The proportion of household of WFH | 38.9% | [13] |
| $nh$ | The cohort size in the school | 15 | [14] |
| $a_r$ | The school attendance ratio of children and youth who will back to school | 0.6834 | [15] |
| $T_1$ | The time of school reopening starts | Sep. 8 | [16] |
| $\Delta T_A$ | Average completing time for all household who send children and youth back to school | 7 | Assumed |
| $\Delta T_q$ | Average completing time for those households staying at home before school reopening who send children and youth back to school | 7 | Assumed |
| $P$ | Total population in Toronto | 2956024 | 0 |
| $n$ | Average number of household population | 2-3 | [2] |
| $N_H$ | The total number of households in Toronto | 1239995 | Calculated |
| $t_s$ | The starting time of school at day t | - | |
| $g_A(t)$ | The daily returning rate of children and youth back to school | - | |
| $G_A(t)$ | The cumulative returning rate of children and youth back to school | - | |
| $g_q(t)$ | The daily returning rate of children and youth from WFH back to school | - | |

| Parameters | Descriptions | Estimated Values | Sources |
| --- | --- | --- | --- |
| $\beta_{gai}$ | The probability of transmission per contact among adults in the community | Jul 31-Sep 8: 2.4100e-02<br>Sep 8-Oct 2: 2.6168e-02<br>Oct 2-Nov2: 1.6868e-02<br>Nov2-Nov23: 2.1868e-02 | Estimated |
| $\beta_q$ | Contact transmission rate within household members | Sep 8-Oct 2: 1.8792e-02<br>Oct 2-Nov23: 0.85e-02 | Estimated |
| $\mu_c$ | Exponential changing rate of contact rate among children and youth in the school | 8.5e-01 | Estimated |
| $c_{aai}$ | The contact rate among adults in the community | Jul 31-Sep 8: 10.364<br>Sep 8-Oct 2: 11.58<br>Oct 2-Nov2: 10.2<br>Nov2-Nov23: 10.564 | Estimated |
| $c_{aci}$ | The contact rate between adults and children in the community | Jul 31-Sep 8: 6.62<br>Sep 8-Oct 2: 7.22<br>Oct 2-Nov2: 3.42<br>Nov2-Nov23: 7.5 | Estimated |
| $c_{cci}$ | The contact rate among children and youth in the community | Jul 31-Sep 8: 9.36<br>Sep 8-Oct 2: 9.5<br>Oct 2-Nov2: 3.8<br>Nov2-Nov23: 5.8 | Estimated |
| $q_{g2}$ | The quarantine rate of symptomatic infection among adults | 6.4214e-01 | Estimated |
| $q_{sc}$ | The quarantine rate of symptomatic infection among children and youth in the school | 9.6996e-01 | Estimated |
| $\eta$ | The efficiency of self-screening procedures for detecting infected children and youth without symptoms | 2.0544e-01 | Estimated |
| $G_q$ | The maximum returning rate of children and youth those in the household working-from-home | 2.5705e-01 | Estimated |

### 3. Data

#### Reproduction numbers and numerical risk assessment

We calculate the reproduction numbers *R_t_* based on the data of daily new cases by episode date of Toronto with different age groups from Jul 31 to Nov 23 and classify the phases as follows, considering the trend of data (*R*_*t*_) and the control measures that public health implemented.

Phase 1: Jul 31 – Sep 7

Phase 2: Sep 8 – Oct 2

Phase 3: Oct 3 – Nov 1

Phase 4: Nov 2 – Nov 23

Also, combined the results of 25 and our estimation in this model, we classify the control measures into three different levels based on the public health reopening stages.

**Figure A1:**
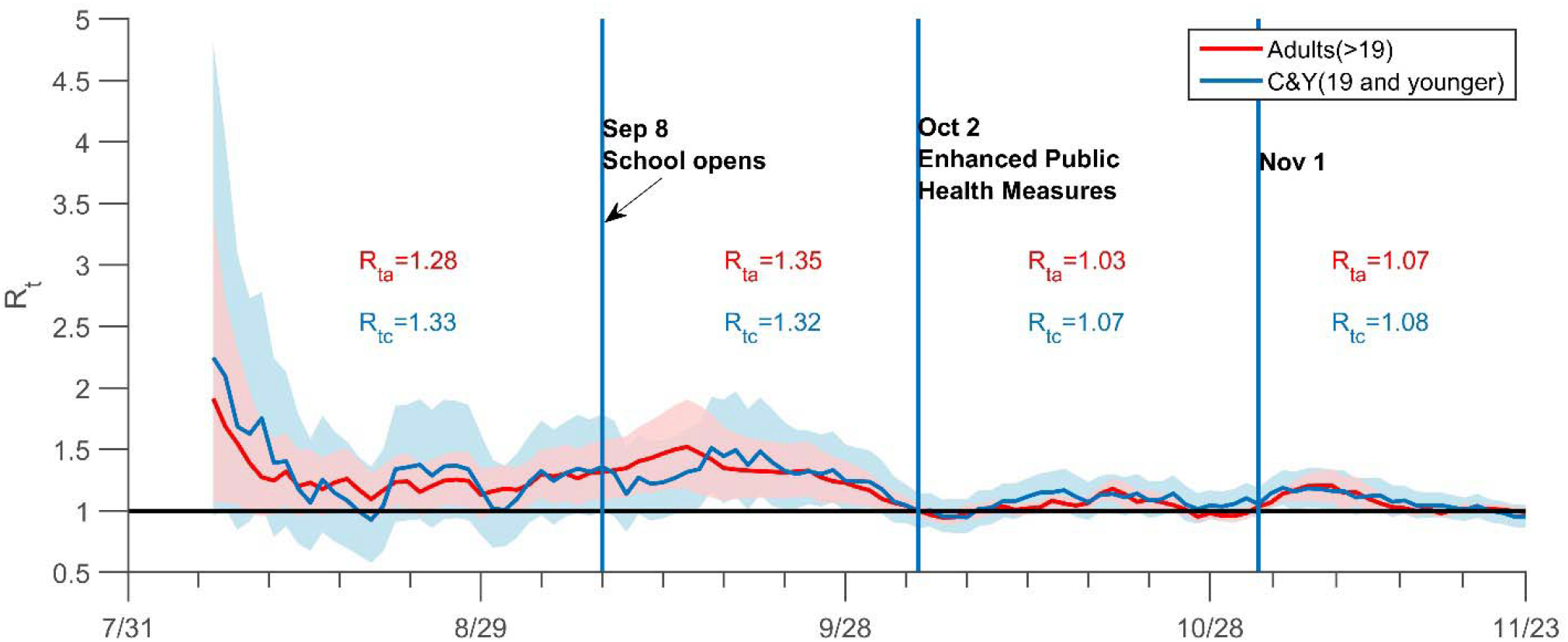
Transmission risk of COVID-19 in Toronto. Instantaneous reproduction number (*R_t_*) in Toronto from Jul 31 to Nov 23. The average *R_t_* Nov 23). The dark solid line indicates the before and after the school reopening (Sep 8), and in phase 3 (Oct 2 -Nov 1) and phase 4 (Nov 1-Nov 23). The dark solid line indicates the critical threshold *R_t_*=1. C&Y is children and youth. All dates are in 2020.

**Table A5.** The summary of different levels of control measures

| Stages | Control measures | The assumed performance of public control measures |  |
| --- | --- | --- | --- |
|  |  | Contact rate among adults in the community | Probability of transmission per contact |
| Stage 1<br>(May 19-June 24) | Open select workplaces, allow some small gatherings. | 8.561 | 0.0168 |
| Stage 2<br>(June 24-July 31) | Open more workplaces and outdoor spaces, allow some larger gatherings. | 9.429 | 0.0168 |
| Stage 3<br>(July 31- October 10) | Further relax the restrictions on public gatherings, opening all workplaces responsibly. | 10.364 | 0.019 |

#### Parameter estimation and data fitting

Using the cumulative confirmed case data of adults and children and youth by episode date in Toronto from Jul 31 to Nov 23, we fit our model by the least-square method to estimate the parameters. The results show that our model fits very well with the Normalized Mean Square Error (NMSE) = 0.995. Parameters are evaluated and presented in Table A4.

**Figure A2:**
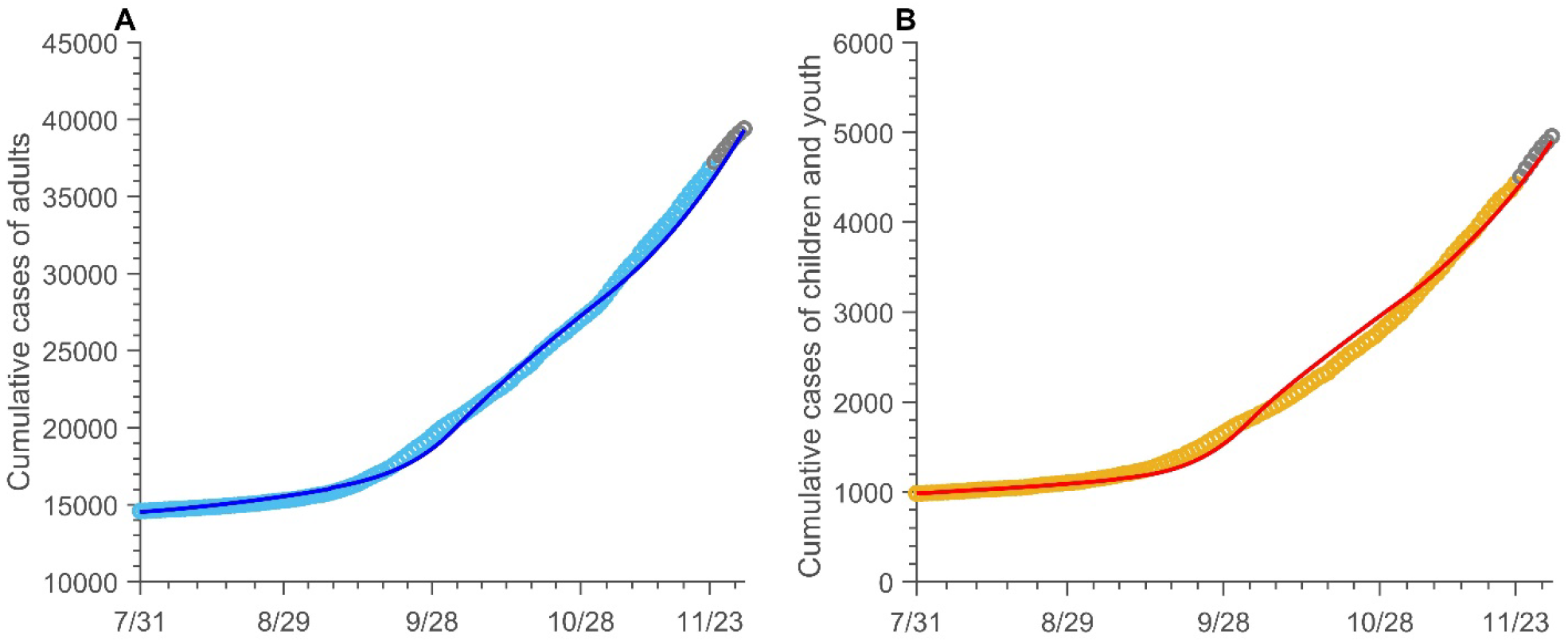
Cumulative COVID-19 incidence of adults, and children and youth in Toronto. Data fitting of COVID-19 infection in Toronto from Jul 31 to Nov 23, 2020. The blue (adults, >19) and yellow (children and youth, ≤19) circles represent real data. The blue (adults, >19) and red (children and youth, ≤19) solid curves are from model simulations. The grey circles are the data for validation from Nov 24 to Nov 30. All dates are in 2020.

### 4. Control the risk in the school

Our findings suggest that school control measures, such as self-screening, limited attendance, reduced class sizes, alternating in-person learning, reduced schooling days are effective in reducing children and youth infections in schools. In-person attendance has the most obvious impact on school infections. If 90% of children attend school in person figure A3B) the daily new cases (reaching 132 in December) was much higher compared to the scenario with only 10% of attendees in school, showing only a maximum of 14 infections per day, dropped by 68.2%. Reducing school opening days had the least impact on the cumulative infections of children and adults by Dec18. When the school opening days are reduced from 5 days to 1 day a week, the numbers of cases among children and adults population only decreased by 2.3% and 0.66%, respectively (table A6). When the cohort size decreases from 30 to 10, the school’s new infections on Dec 18 decreased by 7.5%, and the total cumulative infections of children and adults on Dec 18 decreased by 3.9% and 1.1%, respectively (table A6). However, the difference is not visible when the self-screening efficiency is improved from 10% to 90%.

**Figure A3:**
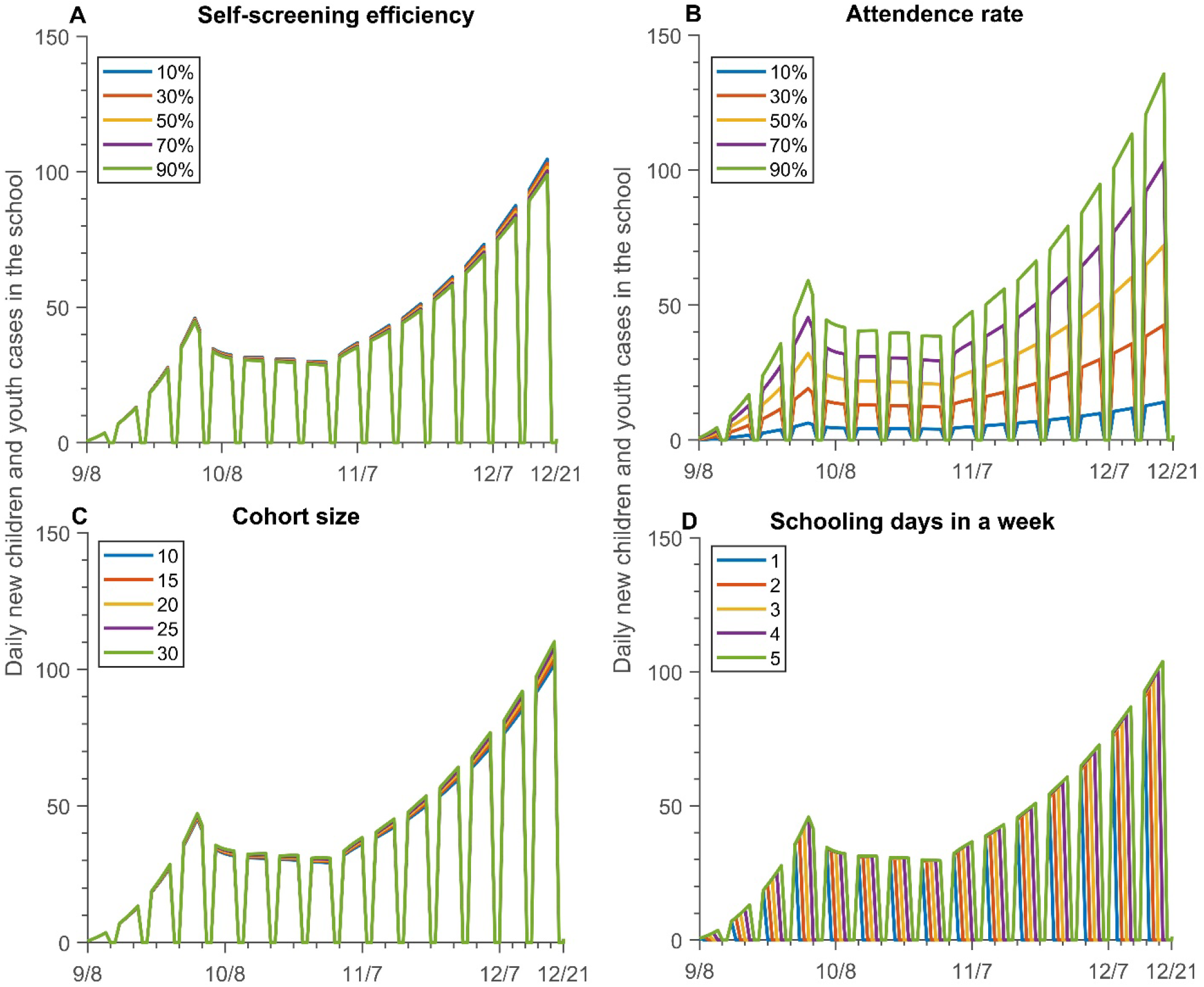
The number of new children and youth’ cases in the school with different mitigation strategies. (A) different level of self-screening measures; (B) different in-person attendance; (C) different cohort sizes; (D) different opening days.

**Table A6:** The symptomatic cases under different school mitigation strategies

| Control measures | Cases | New cases of children and youth in the school |  | Cumulative case of children and youth |  | Cumulative case of adults |  |
| --- | --- | --- | --- | --- | --- | --- | --- |
|  |  | Oct 1 | Dec 18 | Oct 1 | Dec 18 | Oct 1 | Dec 18 |
| Self-screening efficiency | 10% | 43 | 102 | 1678 | 6878 | 19377 | 51637 |
|  | 30% | 42 | 100 | 1676 | 6830 | 19376 | 51536 |
|  | 50% | 42 | 99 | 1674 | 6782 | 19375 | 51435 |
|  | 70% | 42 | 98 | 1672 | 6736 | 19374 | 51336 |
|  | 90% | 41 | 96 | 1671 | 6690 | 19373 | 51237 |
| Attendance rate | 10% | 6 | 14 | 1670 | 6682 | 19373 | 51277 |
|  | 30% | 18 | 42 | 1672 | 6722 | 19374 | 51351 |
|  | 50% | 30 | 70 | 1674 | 6776 | 19375 | 51450 |
|  | 70% | 42 | 100 | 1677 | 6850 | 19376 | 51579 |
|  | 90% | 55 | 132 | 1680 | 6948 | 19378 | 51749 |
| Cohort size | 10 | 42 | 99 | 1674 | 6786 | 19375 | 51443 |
|  | 15 | 42 | 101 | 1677 | 6852 | 19376 | 51584 |
|  | 20 | 43 | 103 | 1679 | 6920 | 19378 | 51726 |
|  | 25 | 43 | 105 | 1682 | 6988 | 19379 | 51871 |
|  | 30 | 44 | 107 | 1684 | 7059 | 19381 | 52018 |
| Schooling days in a week | 1 | 0 | 0 | 1671 | 6695 | 19373 | 51246 |
|  | 2 | 0 | 0 | 1672 | 6734 | 19374 | 51329 |
|  | 3 | 42 | 0 | 1674 | 6775 | 19375 | 51421 |
|  | 4 | 42 | 100 | 1675 | 6813 | 19375 | 51503 |
|  | 5 | 42 | 101 | 1677 | 6852 | 19376 | 51584 |

Note: Oct 1, two weeks after the school full opening on Sep 17. Dec 18, the last date of fall semester.

### 5. What will happen with the new strain virus

We consider the impact of the new strain on the number of cases (figure A4) until May 31, 2021. We assume that the transmission probability of this new strain virus increase by 50%, and the children will be more susceptible than before, with 70% susceptibility of adults. Also, we assume it will be the main strain virus after 1 month of the new strain introduced. We define as baseline the scenario with school closed, community opened under Stage 1 and old strain. We immediately observe that in the worst case scenario (i.e. only the new strain is considered), the cases increases exponentially in both subpopulations. In particular, under the same conditions as the baseline, but with the new strain (figure A4, light blue dashed line), we observe that the numer of case are three times greater than the ones generated by the old strain. If schools are closed, but the community is opened under Stage 2 measures (orange dashed line), the number of cases are 7 times higher. If both schools and community are opened under Stage 2, the increase is slightly higher than 7.5 times.

**Figure A4:**
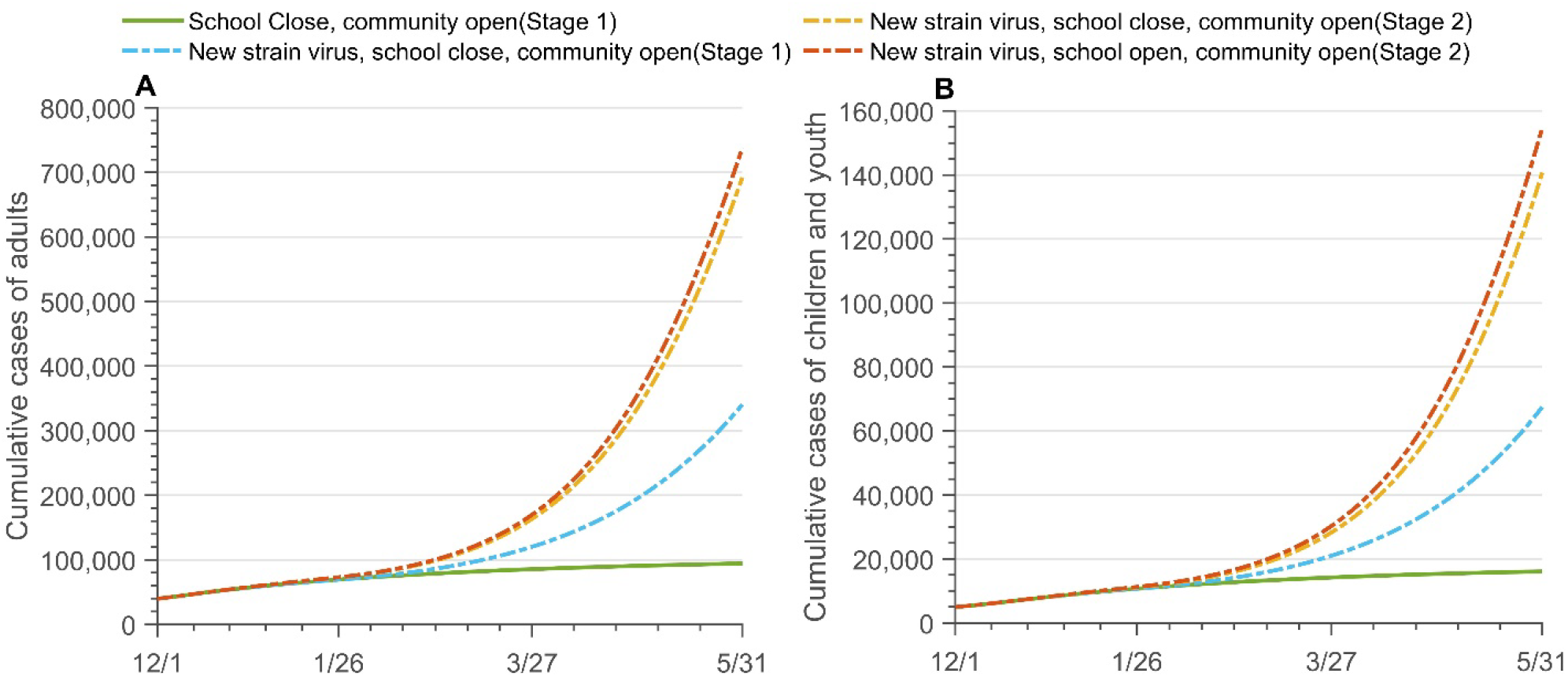
The projection of cumulative cases with different senarioes and new strain. Projection of cumulative cases in adults (A), and children and youth (B) up to May 31, 2021 under different opening strategies, considering the new virus strain, which increases the children’s susceptibility. Green line: old strain, school are closed and community is opened under Stage 1; dashed lines: new strain with closed schools and opened community, in stage 1, (light blue), strain with closed schools and opened community, in stage 2, (orange), strain with opened schools and community, in stage 2, (red).

### 6. Aggressive tracing and testing policy to mitigate

**Figure A5:**
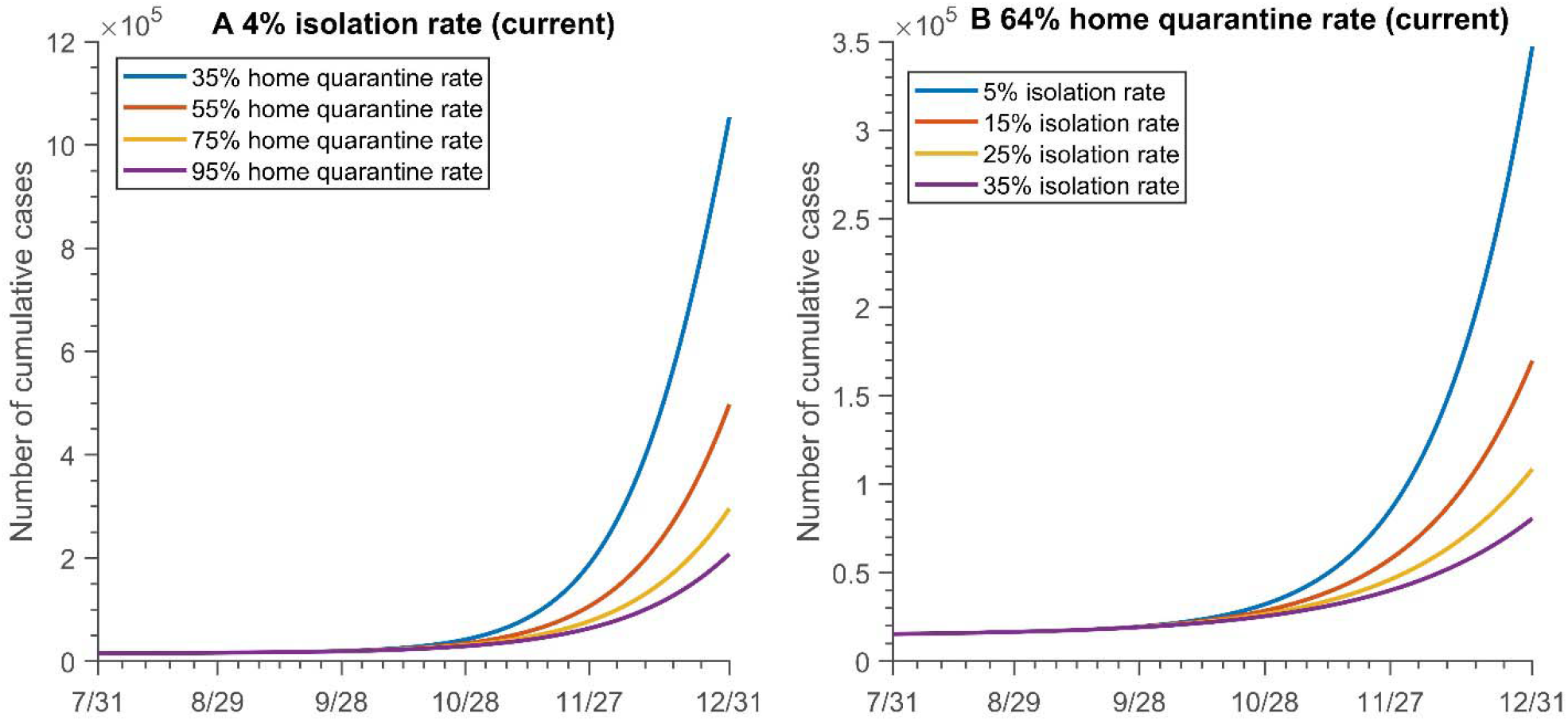
The cumulative cases with varying (A)home quarantine rate and (B) isolation rate.

